# Efficacy and potential human and economic impact of a broadly protective betacoronavirus vaccine

**DOI:** 10.1101/2025.10.17.25338244

**Authors:** George W. Carnell, Robert Vendramelli, Charles Whittaker, Jonathan A. Holbrook, Srivatsan Parthasarathy, Hazel Stewart, Joey Olivier, Bryce Warner, Thang Truong, Charlotte George, Sneha B. Sujit, Sruthika K. Ashokan, Maria Suau Sans, Chloe Qingzhou Huang, David A. Wells, Paul Tonks, Martina Pfranger, Sebastian Einhauser, Patrick Neckermann, Diego Cantoni, Andrew C. Y. Chan, Laura O’Reilly, Luis Ohlendorf, Isaac Unwins, Stefan P. Rautenbach, Matteo Ferrari, Andrew E. Firth, Johannes Geiger, Christian Dohmen, Verena Mummert, Anne Rosalind Samuel, Christian Plank, Joanne Marie M. Del Rosario, Nigel Temperton, Benedikt Asbach, Simon D. W. Frost, Rebecca Kinsley, Sneha Vishwanath, Sofiya Fedosyuk, Ralf Wagner, Darwyn Kobasa, Jonathan L. Heeney

## Abstract

Within two decades, three different betacoronaviruses (β-CoV) have spilled over from animals to humans causing high consequence diseases, the most recent causing the COVID-19 pandemic. Using computational antigen design technology, we developed a single multivalent vaccine candidate capable of broadly neutralising diverse animal and human β-CoVs. We then demonstrated that this single vaccine could protect mice from lethal infection against the three genetically distinct human SARS-CoV, MERS-CoV and SARS-CoV-2 viruses. Epidemiological modelling revealed that a single broadly protective β-CoV vaccine (BPBV) of moderate efficacy (50% against severe disease) could have averted a significant fraction of deaths during the SARS-CoV-2 pandemic, depending on the degree of clinical preparedness established pre-pandemic. Deaths averted by the BPBV varied in our simulations, ranging from 47% if the BPBV had been clinically evaluated through to a phase 2 trail, and ready for vaccine efficacy trials early during an initial outbreak, to 13% if phase 1 clinical testing of such a BPBV had not yet been initiated. Taken together we provide pre-clinical evidence of robust efficacy of a single BPBV vaccine candidate and modelling data demonstrating the significant positive impact on human health and significant economic benefit to national economies which would be achieved by clinically advancing and stockpiling a Phase 3 ready BPBV vaccine in the event of new virus outbreaks.

## Introduction

Emergence of high consequence coronaviruses such as SARS-CoV, SARS-CoV-2, and MERS-CoV highlight the fine balance between human health and zoonotic viral threats^1–3^. While SARS-CoV-2 caused the global COVID-19 pandemic, SARS-CoV and MERS-CoV have caused significant human outbreaks in humans with different transmission dynamics between humans and animals^4^. The COVID-19 pandemic drove dramatic improvements in the timelines of vaccine development, from pre-clinical studies to clinical trials, and further accelerated large scale manufacturing of vaccines for countries with capacity and infrastructure. Within two years, vaccines against SARS-CoV-2 greatly contributed to preventing severe COVID-19 disease and significantly reducing human fatalities ^5–7^.

Advances in vaccine technologies such as mRNA-based prophylactics, have resulted in new vaccine candidates for multiple respiratory virus diseases of humans such as SARS-CoV-2, RSV and seasonal Influenza ^8–10^. However, these vaccines are largely wildtype strain-derived and have reduced efficacy as new variants emerge. The absence of broadly protective vaccines early in the COVID-19 pandemic resulted in waves of variants of SARS-CoV-2 less susceptible to vaccine or infection derived immunity^11^. This underscored two un-addressed problems. First the need for vaccine antigens or approaches that provide breadth of protection from variable groups of viruses, such human relevant coronaviruses (SARS-CoV, SARS-CoV-2 and MERS-CoV), with advances being made in antigen design or combinations of wild type antigens to cover the diversity of these three groups of viruses^12–15^. Second, early implementation of broadly protective vaccines to contain and control potential pandemics before escape mutants evolve. To address these problems, we used a combination of computational, structural and pathogen genome technologies to develop a broad multivalent vaccine candidate. Using an adaptable and rapidly scalable mRNA vaccine platform for early clinical readiness we then validated the efficacy of this vaccine against the diverse group of betacoronaviruses that threaten humans globally.

## Results

Breadth of protection against human variants and animal reservoir viruses was achieved by engineering digitally immune optimised synthetic vaccine (DIOSynVax, DVX) antigen genes that can be expressed in a variety of different vaccine manufacturing platforms including mRNA, in this case delivered as mRNA formulated in ionisable lipidoids (ETHRIS GmbH, Germany). Each antigen component of this DIOSynVax Broadly Protective Betacoronavirus vaccine (DVX-BPBV) was individually selected based on their ability to induce greater immune breadth compared to vaccine candidates based on single wildtype virus antigens used in prototype SARS-CoV-2 vaccines, or to neutralize MERS-CoV related betacoronaviruses including those isolated from camels (Figure 1). We previously reported on the use of this technology to develop a single computational pan-sarbecovirus antigen (S_T2_20)^15^, designed to provide breadth spanning SARS-CoV and SARS-CoV-2 clades (Figure 2B, second panel). To expand the breath across the SARS-CoV-2 variants circulating in humans, we combined this antigen with S_T2_35, a stabilised DVX designed spike-like antigen that alone was able to elicit broadly neutralising antibody responses to SARS-CoV-2 variants of concern (Figure 2B, third panel)^16^. The third component, S_T2_37, is a MERS-CoV spike-based antigen that elicits neutralising antibodies to MERS-CoV and its animal derived camel viruses from Arabian and African viral lineages (Figure 2C, second panel). To recruit pre-existing T-cells and translate to the previously exposed or infected human population we included a broadly reactive pan-sarbecovirus nucleoprotein (NP) antigen, N_T2_2, designed to mimic sarbecovirus nucleoprotein sequences from clades 1a and 1b. This construct induced antibodies and positive T-cell responses measured by ELISpot in our initial immunogenicity studies in mice (Supplementary Figure 1 A-C). These four described components in mRNA were co-formulated into ionisable lipidoids to produce the DVX-BPBV vaccine candidate. This was evaluated for immunogenicity in two different species, mice and guinea pigs, the former with documented comparative neutralising antibody responses to humans^17^. We then tested for vaccine efficacy in pre-clinical trials in mice, with parallel challenges against SARS-CoV, MERS-CoV and SARS-CoV-2 viruses.

**Figure 1.**
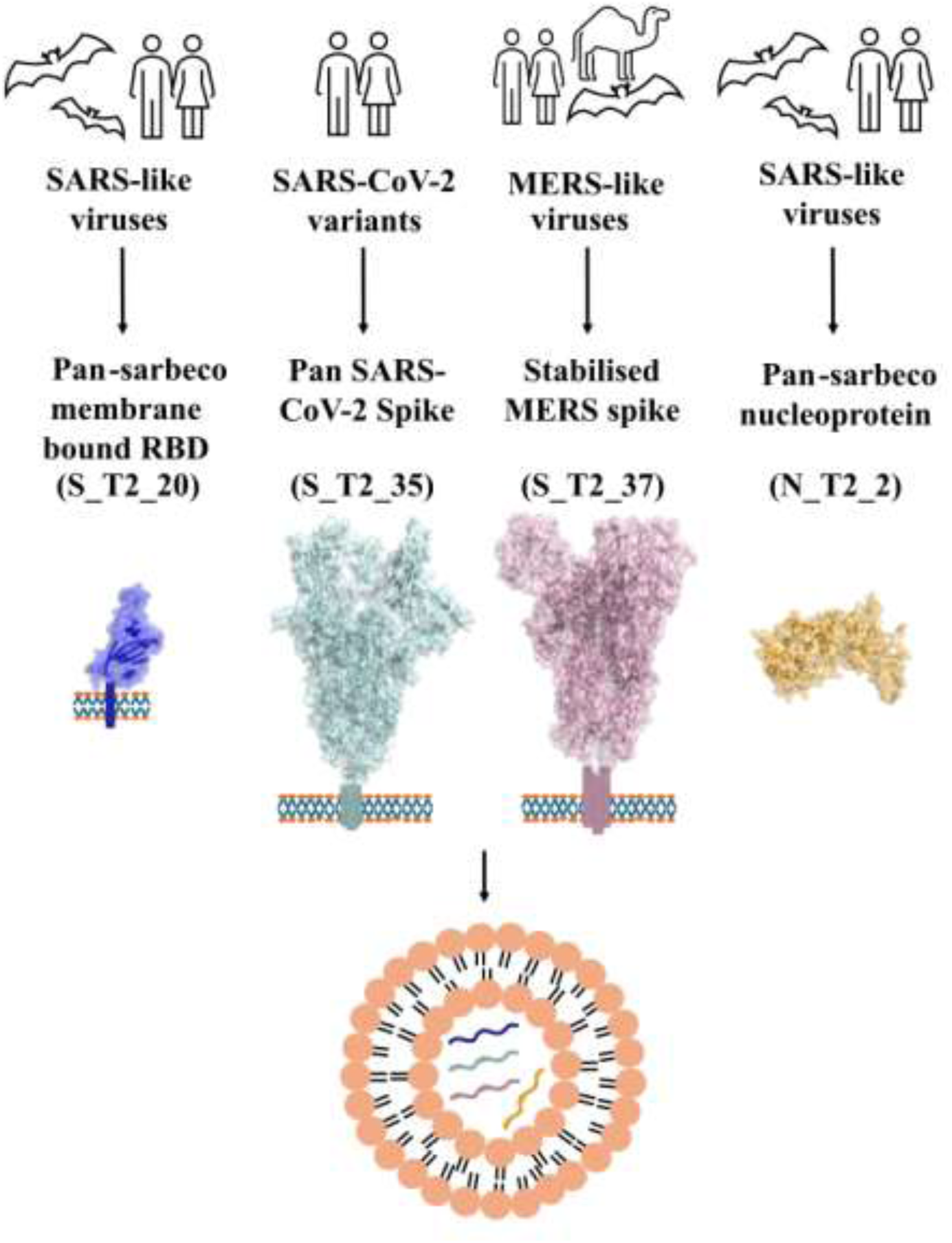
DVX-BPBV approach to a broadly protective pan-betacoronavirus vaccine. To address the threat of multiple viruses with one vaccine, mRNAs encoding four different vaccine antigens, comprising a pan-sarbecovirus membrane bound receptor binding domain (S_T2_20), a pan-SARS-CoV-2 spike antigen (S_T2_35), a stabilised MERS-like spike antigen (S_T2_37) and a pan-sarbecovirus nucleoprotein antigen (N_T2_2), were combined and co-formulated into the same lipid nanoparticle.

**Figure 2.**
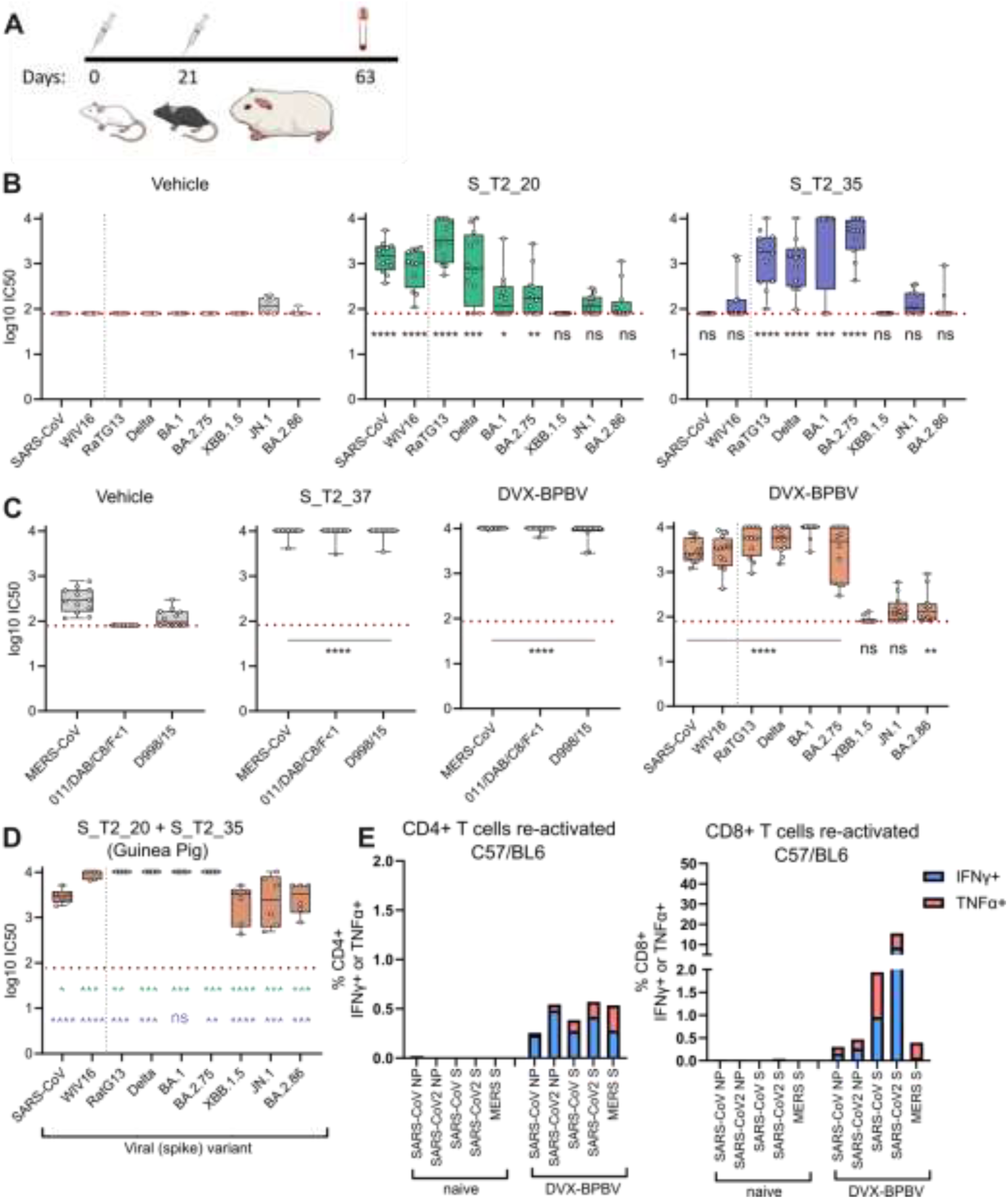
Immunogenicity of single and combined antigens in mice and guinea pigs. A) Animal study schematic for two dose mRNA immunogenicity studies in BALB/c or C57BL/6 mice or guinea pigs. mRNA doses administered at Day 0 and Day 21. Terminal bleed and splenocytes taken at Day 63 for final analysis. B) Pseudotype based neutralisation titres (IC_50_) of vehicle, S_T2_20 and S_T2_35 single antigen groups against a wide panel of sarbecoviruses using Day 63 serum. C) Pseudotype based neutralisation titres of vehicle, S_T2_37 and DVX-BPBV vaccines against the merbecoviruses MERS-COV, African and Arabian lineages (011/DAB/C8/F<1, Kenya and D998/15, United Arab Emirates). Pseudotype based neutralisation of the DVX-BPBV group against the panel of sarbecoviruses matching panel B. D) Pseudotype neutralisation titres from female Hartley guinea pigs immunised with S_T2_20 and S_T2_35 across the broad panel of sarbecovirus spikes, showing enhanced neutralisation of contemporary variants XBB.1.5, JN.1 and BA.2.86. E) Murine splenocyte T-cell flow cytometry analysis from the C57BL/6 DVX-BPBV vaccinated group, CD4+, CD8+, IFNγ and TNFα. All statistical comparisons using two-tailed Mann-Whitney, black asterisks indicate comparisons with the Vehicle only controls. Asterisks in green compare bivalent construct in guinea pigs (D) groups with S_T2_20 in mice, B centre panel, those in blue compare the bivalent vaccine with S_T2_35 in B right panel. ****, *p*<0.0001, ***, *p*≤0.001, **, *p*≤0.01, *, *p*≤0.1.

### Broad neutralising antibodies are elicited by the DVX-BPBV vaccine candidate

We constructed a panel of lentiviral pseudotypes bearing the spike proteins from merbecoviruses and clade 1a/1b sarbecoviruses to assay immune responses generated by our antigens alone, and the DVX-BPBV candidate (Supplementary Figure 2). First, we evaluated the individual S_T2_20 antigen across a panel of SARS-CoV-2 and SARS-CoV viruses (Figure 2B, second panel). Sera from immunised animals neutralised SARS-CoV, WIV16, RaTG13 and Delta (B.1.617.2) using pseudotype virus neutralisation assays as previously described^18^. Neutralising antibody titres induced in S_T2_20 immunised animals were significantly higher than controls against SARS-CoV-2 BA.1 and BA.2.75 but not for later variants (XBB.1.5, 2023, JN.1 and BA.2.86, 2024). We evaluated antibodies elicited by the vaccine antigen S_T2_35, which neutralised variants of concern of SARS-CoV-2, spanning Delta to BA.2.75 with a reduction in titre for XBB.1.5 and later variants JN.1 and BA.2.86 (Figure 2B, third panel)^19^. The S_T2_37 antigen elicited neutralising antibodies to MERS-CoV as well as merbecovirus spikes from closely related viruses of the African (011/DAB/C8/F<1, MK357908.1, Kenya) and Arabian (D998/15, KX108943.1, United Arab Emirates) lineages isolated from camels (Figure 2C, second panel). Vehicle control sera displayed a low nonspecific inhibitory background against MERS-CoV pseudotyped viruses, but significantly lower than positive sera (p=<0.0001) that are at the higher limit of detection of the assay (Figure 2C, first panel). The pan-sarbecovirus nucleoprotein antigen N_T2_2 elicited antibodies to the SARS-CoV-2 nucleoprotein, with no difference between *in vivo* titrated doses between 1 µg and 10 µg per mouse (Supplementary Figure 1A).

In mice, the DVX-BPBV candidate combining each of the above four components elicited strong (average log_10_IC_50_ > 3) neutralising antibodies against the coronavirus spike pseudotype virus panel including SARS-CoV, WIV16, Delta, BA.1 and BA.2.75 where neutralising antibody titres were lower for strains which emerged more recently such as XBB.1.5, JN.1 and BA.2.86 (Figure 2C). Mice immunised with the DVX-BPBV vaccine also elicited strong neutralising antibodies to the merbecovirus pseudotyped viruses tested including MERS-CoV and representative spikes from African and Arabian camel lineages of merbecovirus, extending the neutralising response detected from each mouse to MERS-CoV, SARS-CoV and SARS-CoV-2 clades (Figure 2C). In parallel, we evaluated our sarbecovirus candidates in Hartley guinea pigs, combining the S_T2_20 and S_T2_35 antigens to elicit a pan-sarbecovirus neutralising antibody spectrum. Following two immunisations at 21-day intervals, matched with our studies in mice, the terminal sera revealed even greater titres and breadth of neutralisation, a distinct difference to monovalent data from mice. The antibodies elicited were able to neutralise the entire pseudotype virus panel including the contemporary SARS-CoV-2 variants XBB.1.5, JN.1 and BA.2.86 (Figure 2D). Statistical group comparisons for neutralising antibody are shown in Supplementary Tables 1, 2 and 3.

### T-cell responses elicited by DVX-BPBV

The antigens S_T2_20, S_T2_35 and S_T2_37 were designed to elicit neutralising antibodies to relevant sarbecoviruses and merbecoviruses of human relevance. However, we evaluated T-cell responses elicited by single-antigen or DVX-BPBV vaccines that could contribute to protection from infection or disease. Immunisation of mice with the T-cell antigen N_T2_2 alone provided us with ELISpot (IFNγ) T-cell data (Supplementary Figure 1B and C) which indicated an increased T-cell response compared to wild type nucleoprotein in the same platform (N_T1_1, SARS-CoV-2 NP, Wuhan isolate) though not reaching statistical significance. This trend was present when splenocytes were activated by peptides for both the nucleoproteins from SARS-CoV and SARS-CoV-2, indicating superior expression or stability of N_T2_2 over the matched control antigen N_T1_1, as N_T2_2 elicited splenocytes responded higher than N_T1_1 stimulated by its homologous peptide pool. To characterise the cellular immune responses in more detail, we employed the Flow Cytometry-based intracellular cytokine staining method. This approach evaluates separate activation of CD4+ or CD8+ arms of immunity by quantifying IFNγ and TNFα production in isolated splenocytes from immunised mice following their re-exposure to antigen-specific peptide pools. In addition to the canonical C57BL/6 mouse strain with Th1 type predominant responses, we also tested cellular immune responses in BALB/c mice that are typically used to test humoral immune responses and feature predominant Th2 activation. All tested peptide pools (SARS-CoV NP, SARS-CoV-2 NP, SARS-CoV S, SARS-CoV-2 S, MERS S) re-activated splenocytes isolated from either C57BL/6 (Figure 2E) or BALB/c mice (Supplementary Figure 3 A-D) immunised with DVX-BPBV. Both CD4+ and CD8+ arms of cellular immunity were activated for all antigens to similar extents except for the MERS S CD8+IFNγ response. For SARS-CoV and SARS-CoV-2 S pools, CD8+ responses were 4 and 10-fold stronger than CD4+ responses, respectively (Figure 2E). Differences in cytokine predominance were observed, where splenocytes from BALB/c mice produced significantly more TNFα and responses in splenocytes from C57BL/6 animals was driven mainly by IFNγ. In the BALB/c model, responses in the DVX-BPBV vaccinated group were weaker when compared to respective single-antigen controls (Supplementary Figure 3A, 3C), for example S_T2_35 vs DVX-BPBV, however, in C57BL/6 the opposite was observed with overall strongest cytokine production in DVX-BPBV immunised mice (Supplementary Figure 3B, 3D).

### DVX-BPBV protects mice against SARS-CoV, SARS-CoV-2 and MERS-CoV

To evaluate the protective effect of our measured immune responses, we carried out three parallel vaccine challenge studies to confirm if the immune responses correlated with vaccine efficacy against live virus challenge in susceptible mice. We challenged vaccinated mice with SARS-CoV, SARS-CoV-2 (Delta, B.1.617.2) and MERS-CoV as prototype viruses from the three human Coronaviruses with spill overs occurring between 2003 and 2019. The SARS-CoV-2 Delta variant was chosen as it was the most recent available variant causing lethal infection in our mouse model. Mice were immunised twice with a 21-day interval, bloods sampled as indicated in Figure 3A, and animals challenged on Day 42, 21 days after the second vaccine dose. Details of the viruses and animals used in this study are listed in Figure 3B and Supplementary Table 4 and 5.

**Figure 3.**
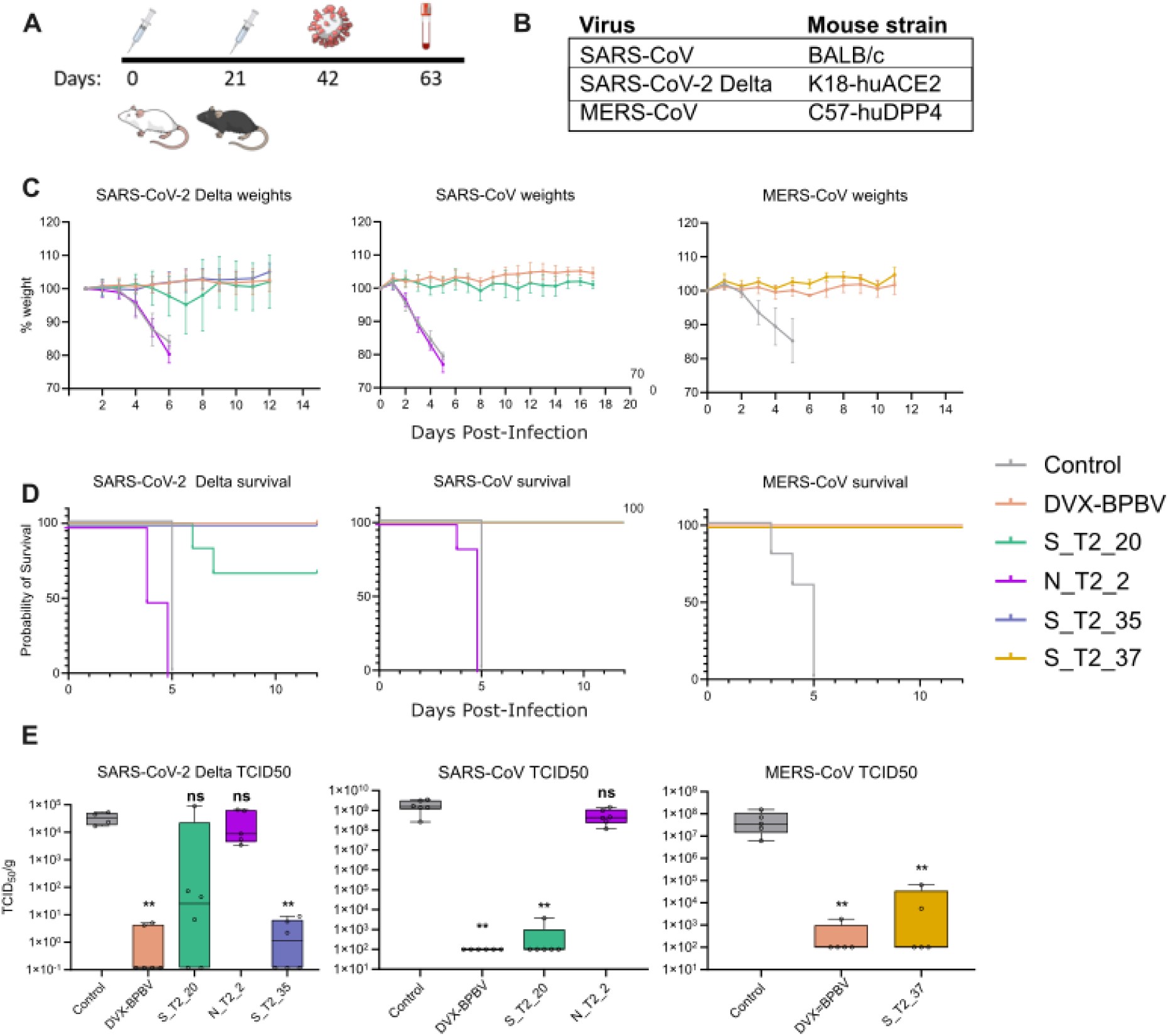
Preclinical efficacy of single and DVX-BPBV vaccines in a murine lethal challenge experiment. A) Animal study schematic for two dose mRNA immunogenicity followed by lethal challenge with (B) mouse adapted SARS-CoV in BALB/c mice, SARS-CoV-2 (Delta, B.1.617.2) in K18-huACE2 mice, or MERS-CoV in C57/huDPP4 mice. C) Percentage weight loss of vaccinated, challenged mice weighed daily after infection with SARS-CoV-2 (left), SARS-CoV (centre) or MERS-CoV (right) (D) Probability of survival analysis of challenged mice over a 14-day window, showing 100% of survival for the DVX-BPBV group challenged with SARS-CoV-2 Delta (left), SARS-CoV (centre) and MERS-CoV (right). E) SARS-CoV-2 (left), SARS-CoV (centre) and MERS-CoV (right) viral lung titre (TCID50) measured from a subset (n=6 per group) of homogenised lungs taken from vaccinated and challenged mice on D3 post infection. All statistical comparisons using two-tailed Mann-Whitney U test, asterisks denote significance compared to the vehicle only group. **, *p*≤0.01, *, *p*≤0.1.

Mice immunised with the DVX-BPBV vaccine did not display clinical symptoms or show weight loss (Figure 3C), and 100% of mice survived infection from all three viruses (Figure 3D). Virus was negligible or not detectable in the lungs at day 3 post infection, in contrast to naïve mice (Figure 3E). In single antigen positive controls, the spike-based antigens alone afforded a degree of protection; the S_T2_20 antigen protected 100% of mice from SARS-CoV but only 66% of mice from SARS-CoV-2 Delta (Figure 3C, left panel) while the broad-variant SARS-CoV-2 antigen S_T2_35, alone demonstrated 100% protection of mice from SARS-CoV-2 Delta. Similarly, S_T2_37 protected 100% of mice from MERS-CoV. The internal nucleoprotein N_T2_2 had no positive impact on the prevention of infection or clinical symptoms from SARS-CoV or SARS-CoV-2 Delta as a single antigen (Figure 3C).

### Evidence for public health impact of early availability of broadly protective vaccines

The advent of powerful technologies such as the mRNA vaccine platform have strengthened our ability to react to emerging viruses of medical importance, which will likely continue to spill over from animal reservoirs in the future as human and animal populations overlap. The availability of broadly protective vaccines with coverage spanning multiple betacoronavirus clades of predicted pandemic potential saves critical time to contain and control a potential pandemic, and to avert human disease, social, health care and economic consequences whenever a new pandemic emerges. The effectiveness of such broadly protective vaccines depends on their availability for phase 3 efficacy trials.

We explored the public-health impact that a stockpiled BPBV could have had on COVID-19 mortality in the pandemic’s first year using a previously published SARS-CoV-2 transmission model calibrated to country-specific excess mortality data ^14,15,16^ and evaluated potential BPBV impact using these calibrated fits as a counterfactual. These models assume that the decision around BPBV stockpile activation was made following 100 globally reported COVID-19 deaths whilst exploring 3 scenarios varying the delay between stockpile activation and widespread administration to eligible groups (assumed here to be all individuals aged 60+ years). The first model assumes the BPBV has been fully clinically evaluated ahead of the pandemic and is therefore available for immediate widespread administration; the second model assumes that only Phase 3 trials are required (leading to a delay of 140 days based on data from COVID-19 vaccine trial timings); and the third model assumes minimal clinical evaluation had been conducted ahead of the pandemic, such that evaluation must begin with Phase 1 trials (leading to a delay of 250 days based on data from COVID-19 vaccine trial timings). In all cases, we assume that once available, the BPBV was administered at a vaccination rate specific to World Bank income groups (derived empirically from Our World In Data COVID-19 vaccination data^23^). We also vary assumptions about BPBV efficacy against severe disease (50%, 75%, 95%) and infection (0%, 35%), as well as assumptions about the BPBV stockpile size (and thus the coverage of 60+ year olds that can be achieved). This ranged from 30% (“Low” coverage) to 90% (“High” coverage) and an additional scenario was explored in which the size of the stockpile maintained varied by country (“Variable” coverage) in a manner determined by its World Bank Income Group.

Our results suggest that a BPBV with 75% efficacy against severe disease, 35% efficacy against infection and a stockpile sufficient to vaccinate 50% of the global eligible population which was available immediately for administration could have averted 55% (38%-69%) of COVID-19 deaths during the pandemic’s first year (Figure 4A, 4B & 4E). Impact was reduced if the BPBV first has to undergo clinical evaluation: 34% (12%-51%) (Figure 4A & 4D) in the scenario where only Phase 3 trials are required, and only 15% (0%-23%) (Figure 4A & 4C) in instances where no clinical evaluation is taken ahead of the pandemic (Figure 4F). Increasing stockpile size, efficacy against severe disease and efficacy against infection all increased overall impact (Supplementary Figure 4) though the timeliness of BPBV availability (shaped by the extent of clinical evaluation pre-pandemic) was the single biggest driver of impact in the scenarios analysed here, both globally (Figure 4G & Supplementary Figure 5) and at a country-specific level, as illustrated by results for Canada (Figure 4H), Ecuador (Figure 4I), France (Figure 4J) and Indonesia (Figure 4K).

**Figure 4.**
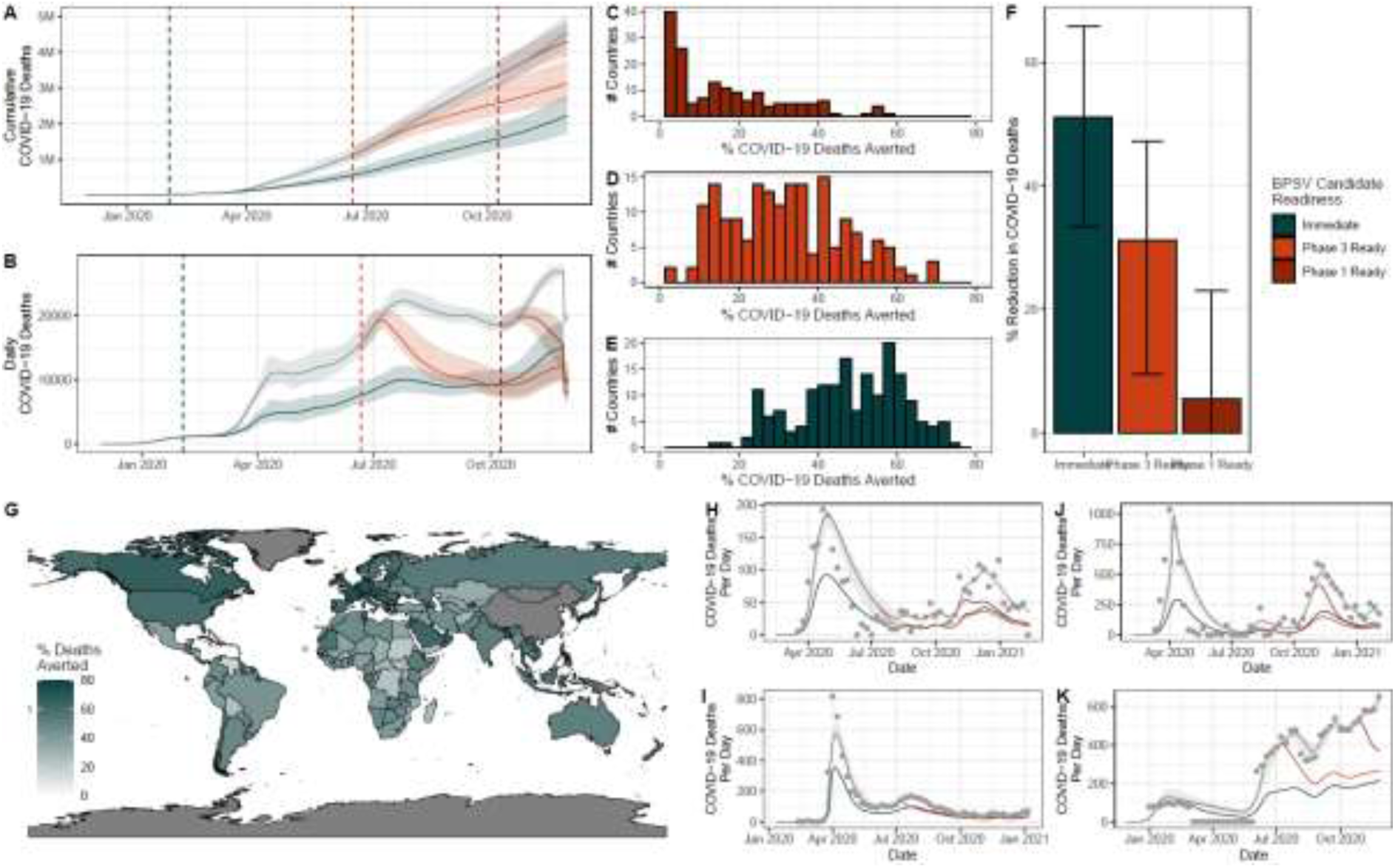
Modelling potential BPBV impact during the COVID-19 pandemic. A) Cumulative global COVID-19 deaths during the first year of the pandemic without (grey) the BPBV, and with the BPBV (lines according to readiness scenario). Immediate readiness = BPBV stockpile activated and administered following 100 globally reported COVID-19 deaths; Phase 3 ready = BPBV evaluated in Phase 3 trials prior to outbreak, delaying administration by 140 days relative to the immediate scenario; Phase 1 ready: no pre-existing BPBV evaluation necessitates starting with Phase 1 trials, leading to further delays relative to the immediate scenario (250 days). B) As for (A) but for daily COVID-19 deaths. C) Histogram of % of COVID-19 deaths averted by the BPBV in each country, for the Phase 1 readiness scenario. D) as for (C), but for the Phase 3 readiness scenario. E) As for (C) but for the Immediate readiness scenario. F) Total COVID-19 deaths averted during the first year of the pandemic by the BPBV for each readiness scenario. Error bars indicate the 95% confidence interval across all countries. G) Modelled impact of the BPSV during the first year of the COVID-19 pandemic in different countries around the world, assuming the immediate readiness scenario. Colour indicates the percentage of COVID-19 deaths in first year of pandemic averted if BPBV is available. H) BPSV impact on COVID-19 mortality in Canada during the first year of its COVID-19 epidemic. Grey line indicates model fit to COVID-19 excess mortality data (light grey points) and ribbon indicates the 95% CI across 100 model simulations using different posterior draws for the Rt trajectory. Coloured line indicates expected mortality when the BPBV is available, with the mean trajectory across 100 simulations plotted. Line colours reflect readiness scenario assumed. I) as for (H) but for Ecuador instead of Canada. J) As for (H), but for France. K) As for (H) but for Indonesia.

A broadly protective betacoronavirus vaccine could have a substantial positive economic impact by averting the need for extended lockdowns. In 2020, the Global GDP contracted 2.7%, compared to the average annual growth of 3.4%, resulting in an estimated loss from the COVID-19 pandemic of ∼$8.5 trillion^24^. In our “Immediate” scenario, the ∼6% lost growth in global GDP (∼$12.4 trillion in 2025) could be averted, while having it ready for Phase I or Phase III, respectively could avert ∼1-2% ($2.06–4.14 trillion) (Supplementary Figure 6). These trillion-dollar savings far outweigh the ∼$900 million cost required to bring a vaccine to market^25,26^.

## Discussion

As the global population expands past 8 billion and humans and their domestic animals continue to encroach with increasing exposure to wildlife habitats and pathogens, spill over events of high consequence emerging and re-emerging pathogens will continue. As the world has recently witnessed with the 3 different betacoronaviruses since 2002, the increased convergence of ecological, agricultural and demographic pressures makes zoonotic outbreaks with pandemic scenarios increasingly likely. We have recently reported the use of computational methods to design novel vaccine antigens mimicking those from a variety of pathogens, including coronaviruses, and demonstrated that they induce antibodies with significantly greater breadth of neutralisation than traditional vaccines ^15,16^. With respect to coronaviruses, the protective effects that we observe in vaccinated mice suggest a significant potential for human vaccines against not only current variants of SARS-CoV-2 but also related viral threats of pandemic potential. Our computationally designed antigens induce broadly neutralising immune responses against multiple members of the betacoronaviruses, such as the emerging and re-emerging SARS-CoV related viruses, with synergistic immune responses in larger animal models (Figure 2D). In mice, this DVX-BPBV vaccine candidate not only prevented viral replication in the lungs but also abrogated clinical symptoms and mortality (Figure 3C, 3D, 3E). Exploiting the scalable advances of the mRNA platform, here we formulated a multivalent combination of validated broadly reactive β-CoV vaccine DVX-antigens into one vaccine payload, maintaining or increasing neutralising antibody potency with no evidence of immunological competition.

We have demonstrated that this single BPBV vaccine candidate elicited protection in parallel against three human betacoronavirus pathogens, specifically SARS-CoV, SARS-CoV-2 and MERS-CoV. All naïve control animals developed fatal disease in the first week of challenge, while immunised animals were protected from all three viruses (Figure 3D). This system is amenable to rapid change using panels of pre-prepared DVX-antigens and provides a future-proof alternative system to pre-empt the next pandemic. These findings provide evidence that next generation vaccines, based on the DVX vaccine antigen technology, could not only provide wide-ranging protection from diverse betacoronaviruses in the event of future zoonotic betacoronavirus spill over events, but can also be applied to vaccines to other zoonotic threats such as influenza virus spill overs from animals. While other betacoronavirus vaccines have been proposed based on combinations of strain specific RBD proteins, or chimeric RBD-spike proteins, none have used in silico designed antigens and tested these through immunogenicity and efficacy studies against all 3 prototype high consequence human coronaviruses^8,9,14,27^.

Our modelling data revealed further evidence of the significant potential impact that a stockpiled BPBV could have during a future beta-coronavirus pandemic. Crucial to achieving this impact however will be comprehensive clinical evaluation of potential candidates ahead of the pandemic. Such evaluation will in turn enable rapid and timely deployment of vaccines during said pandemic, which our analyses highlight as the primary driver of BPBV impact. Experiences with COVID-19 have highlighted the importance in reducing vaccine response timelines, ideally further than the those achieved during the COVID-19 pandemic^28^. However, despite renewed initiatives, recent analyses have suggested that traditional vaccine pipelines are unlikely to be achieved in less than 220 days^28^. The stage of clinical development was found to be the bottleneck. For instance, in the case of a vaccine candidate that had not yet undergone Phase 1 trials would have minimal impact upon public health, in the event of an outbreak of a highly transmissible pathogen, there would be a reduction in public health impact. In contrast an immediately available broad-spectrum vaccine of potential spill over viruses could therefore offer substantial value as an early response at the location of an outbreak to contain and control, as well as providing vulnerable risk-groups with protection. This would buy valuable time before more tailored candidates are developed – but only if the BPBV itself is available and clinically ready to be deployed in an outbreak scenario efficacy trial rapidly following detection of the novel betacoronavirus.

Evaluating BPBV vaccine candidates against animal viruses that have yet to emerge in the human population is likely to prove challenging. Without human transmission and adaptation, there is limited opportunity for pre-clinical evaluation based on classical clinical efficacy end-points. Our work highlights the importance of surmounting this challenge, however there is much that can be done. Further identification and characterisation of robust immune correlates of protection in existing betacoronaviruses (such as the neutralising antibody titres characterised by Khoury et al for SARS-CoV-2^29^) can inform neutralising antibody thresholds, allowing their use as surrogates for clinical protection and supporting immunobridging across platforms and viruses. The FDA’s accelerated approval of the live attenuated chikungunya vaccine IXCHIQ on the basis of a micro plaque reduction neutralisation test, µPRNT-50 ≥ 150 titre, demonstrates the feasibility of such an approach being used to support vaccine approval^30,31^. Additionally, efforts around creating interconnected and centralised laboratory/clinical trial networks, coupled with recent funding announcements supporting computational approaches to support trial design and development of adaptive, statistically efficient trial protocols ahead of time will enable the rapid activation of harmonised trial sites and deployment of regulator ready study designs within days of an emerging virus alert. Together, these developments are likely to greatly expedite the time to evaluation in instances where Phase 3 efficacy trials cannot be completed pre-pandemic. In conclusion new vaccine technologies that provide breadth across groups of pathogens with zoonotic and pandemic potential, such as DVX-BPBV, can play an important role in mitigating future pandemics if they are developed to early-stage clinical readiness for efficacy trials as soon as nascent outbreaks are identified.

## Data Availability

All data produced in the present study are available upon reasonable request to the authors

## Acknowledgements

The authors thank Professor Kei Sato for the JN.1 spike expression plasmid. We also thank <u>NIAID</u> <u>Visual & Medical Arts</u> for the figure art BIOART-000279, BIOART-000196, BIOART-000464, BIOART-000052, BIOART-000506 used in Figures 2 and 3. IU and SPR would like to acknowledge Airfinity Nexa.

## Funding

Coalition for Epidemic Preparedness Innovations (CEPI), broadly protective coronavirus vaccines (GWC, JH, SO, JO, CLG, SBS, MSS, DW, JG, CD, VM, ARS, CP, LO, MF, JMD, RK, SV, SF, JLH).

UKRI/MRC code number MC_UU_00034/1 (DC)

Bavarian States Ministry of Science and Arts (Grant FORCOVID II) (MP, SE, PN, RW)

The National Microbiology Laboratory (Winnipeg, Canada) was also financially supported by the Public Health Agency of Canada. (RV, BW, TT, DW)

Wellcome Trust Senior Research Fellowships (106207/Z/14/Z, 220814/Z/20/Z) (AEF). European Research Council grant (646891) (AEF, HS)

## Author contributions

Conceptualization: GWC, CW, SV, RW, JLH

Methodology: GWC, RV, CW, JAH, HS, BW, DW, IU, SPR, DC, CP, SDWF, SV, SF, DK

Investigation: GWC, RV, CW, JAH, SP, HS, JO, BW, TT, CLG, SBS, SKA, MSS, CQH, PT, MP, SW, PN, DC, AC, LOR, LO, IU, SPR, MF, JMMDR, SF

Visualization: GWC, CW, SAH, DC, SV, SF

Funding acquisition: RW, RK, JLH

Project administration: JLH, RK, DK, RW

Supervision: GWC, SF, RW, AEF, CP

Writing – original draft: GWC, RV, CW, SV, SF, DK, JLH

Writing – review & editing: All authors

## Competing interests

GWC, SP, SBS, SKA, DW, MF, JMD, SF, RK, SV, SDWF, RW, JLH are current or past employees, shareholders or affiliated with DIOSynVax Ltd, Cambridge, United Kingdom. JG, CD, VM, ARS, and CP are current or past employees, shareholders or affiliated with Ethris GmbH, Munich, Germany. SDWF is an employee of Microsoft. IU and SPR are employees of Airfinity Ltd. JLH, GWC, SV, DW, MF are listed as inventors on coronavirus vaccine patents applications US20240285751A1, EP4126033A2, AU2021249525A1 and CA3179038A1. JLH, GWC, SV, DW, MF, BA, MP, PN and RW are listed as inventors on coronavirus vaccine patent application EP4412645A1. CP and CD are listed as inventors on lipidoid patents EP3013964B1, AU2014300980B2, CA2916800C, CN105579584B, IN369064, JP6609246B2, KR102285326B1, US12064484B2 and ZA201509111B.

The remaining authors declare no conflicts of interest.

## Data and materials availability

Data are available from the corresponding author JLH upon reasonable request.

## License information

Creative Commons Attribution 4.0 International (CC BY).

---

**Supplementary figure 1.**
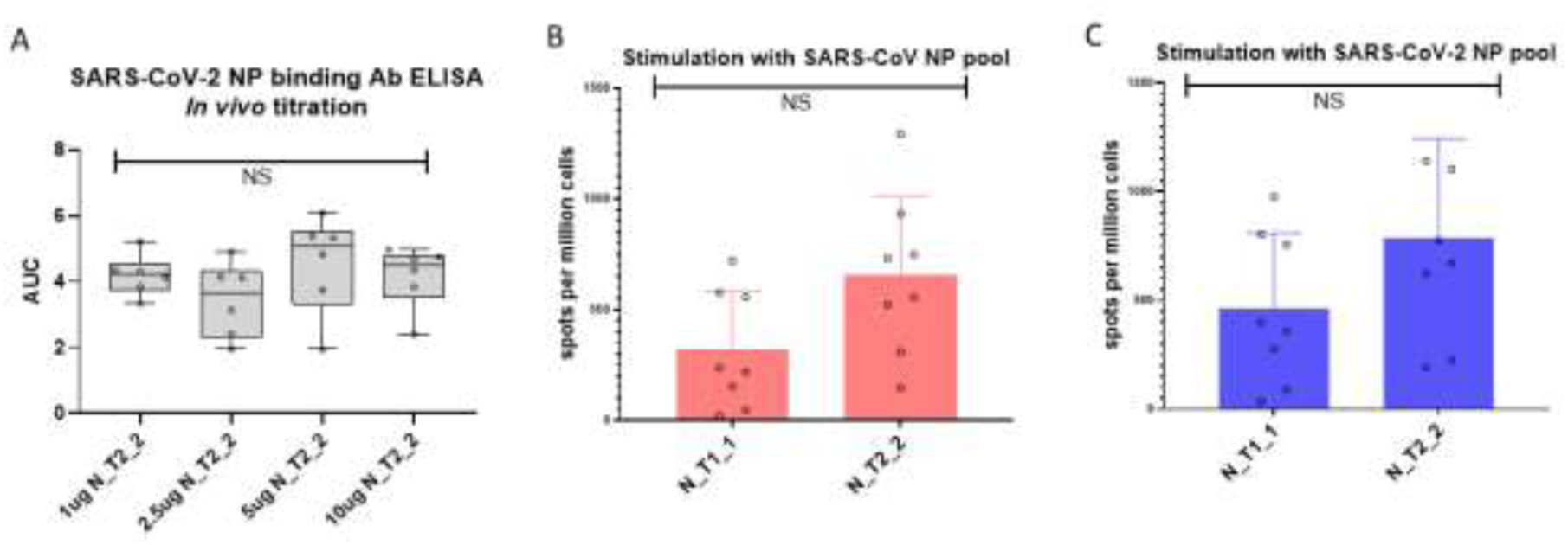
Immunogenicity data for mRNA pan-sarbecovirus nucleoprotein antigen N_T2_2. A) SARS-CoV-2 Wuhan spike direct ELISA binding analysis of the serum generated by an *in vivo* titration of the N_T2_2 mRNA vaccine in BALB/c mice, with mice (n=6 per group) receiving 1, 2.5, 5 or 10µg of mRNA_N_T2_2. Area under the curve for 8-point titration of serum shown. B) and (C), IFNγ ELISpot analysis of BALB/c mice immunised with N_T2_2 or SARS-CoV-2 nucleoprotein (N_T1_1) mRNA (10µg) stimulated with peptide pools from SARS-CoV NP (red) or SARS-CoV-2 NP (blue). All statistical comparisons using two-tailed Mann-Whitney, NS = not significant.

**Supplementary figure 2.**
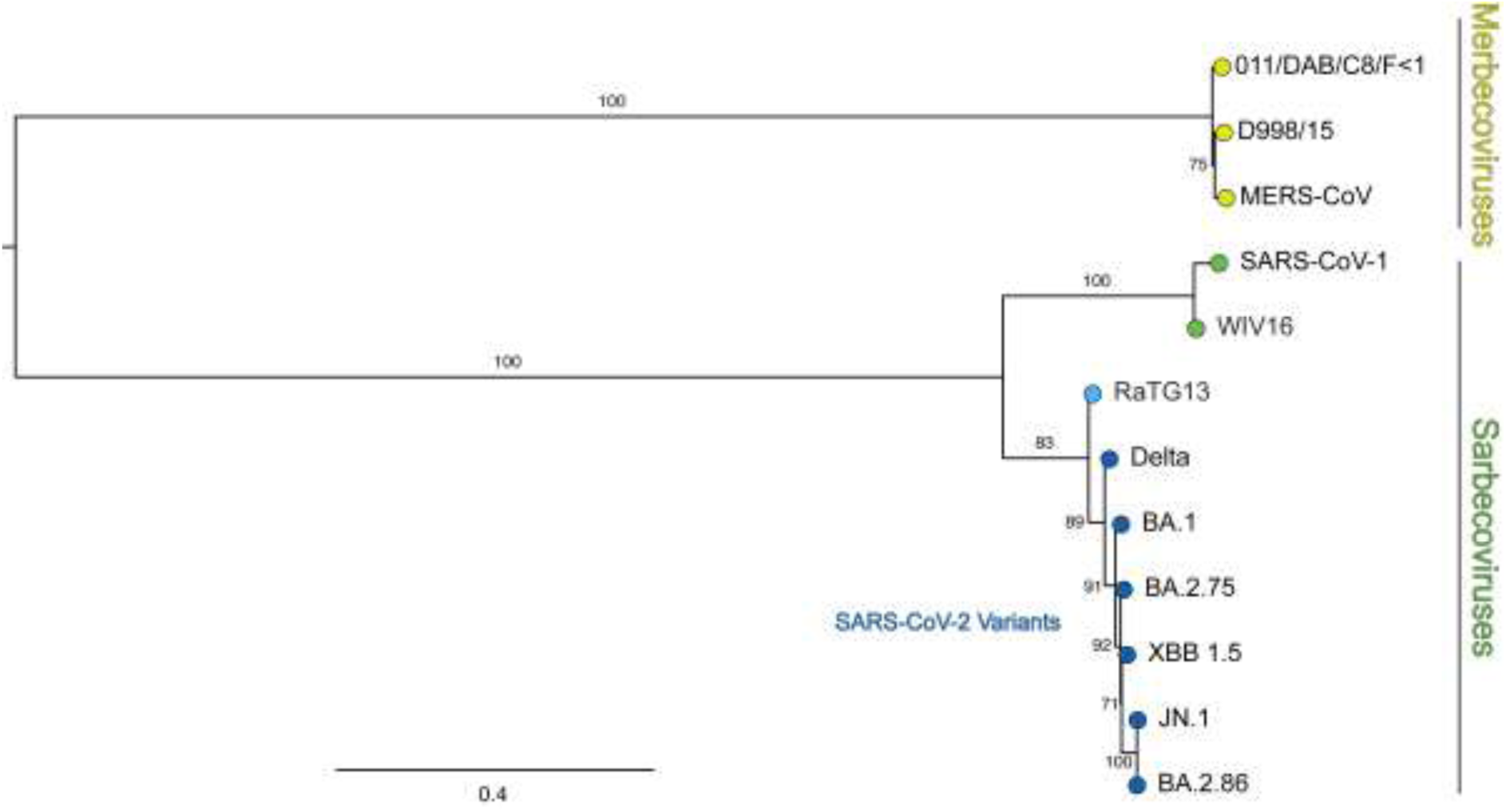
Phylogeny of the Spike protein amino acid sequences used to assess the broadly protective neutralising antibodies from immunised animals. Amino acid sequences representing the spike proteins used in this study were taken from Genbank to produce a maximum likelihood tree, using MAFFT v7 with auto algorithm to align sequences, of which gaps in the alignments were trimmed using TrimAl, and the resultant tree built using IQ-TREE2 with automatic model selection with 1000 ultrafast bootstrap replicated to assess branch support.

**Supplementary figure 3.**
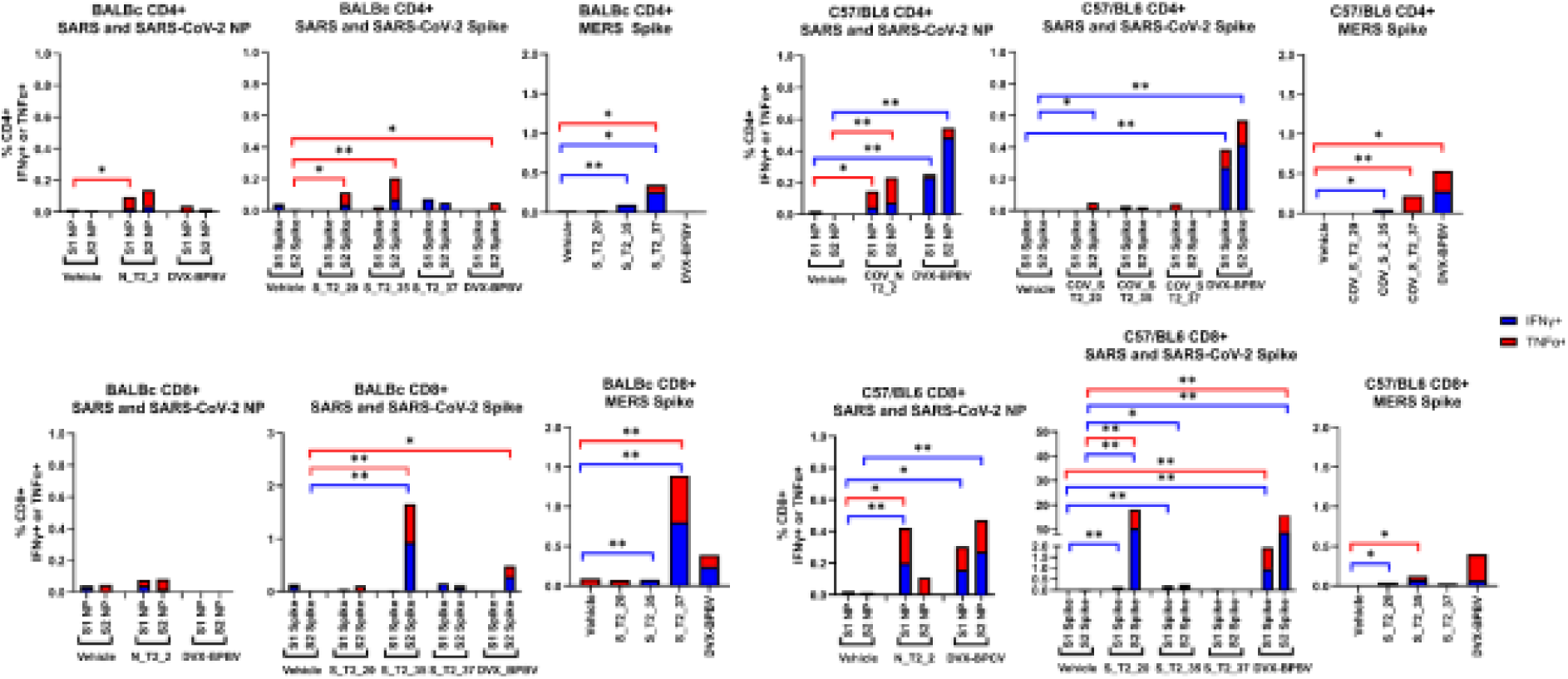
Flow cytometry-based T-cell analysis from BALB/c or C57BL/6 mice immunised single antigen mRNAs. A) BALB/c CD4+ responses against spike or nucleoprotein peptide pools from SARS-CoV, SARS-CoV-2 or MERS-CoV. B) CD4+ responses from vaccinated and naïve (vehicle) immunised C57BL/6. C) CD8+ responses from vaccinated and naïve immunised BALB/c and (D) CD8+ responses from vaccinated and naïve immunised C57BL/6.

**Supplementary figure 4:**
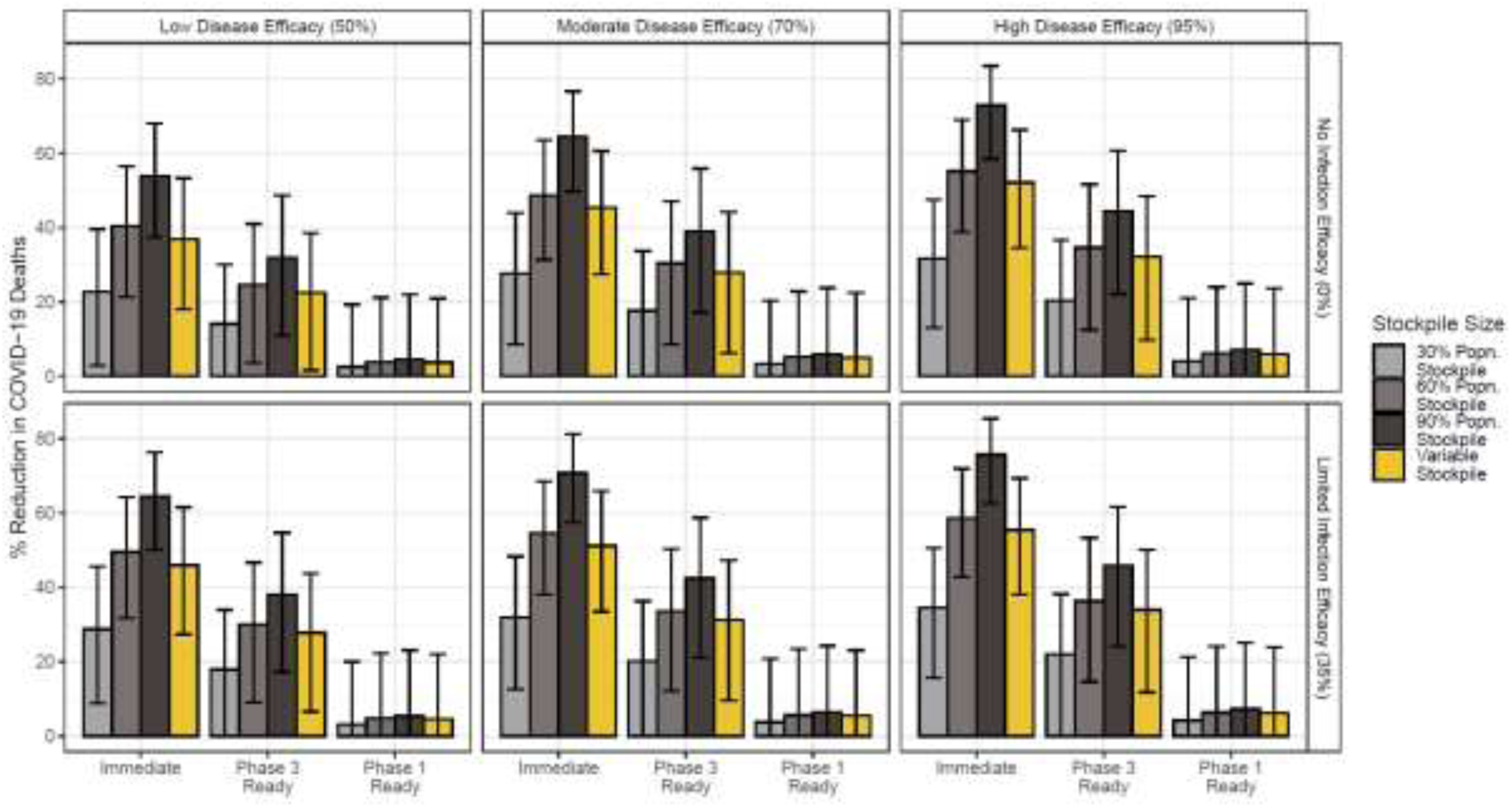
Sensitivity analysis exploring the drivers of BPBV impact. A comprehensive suite of sensitivity analyses varying readiness scenario, BPBV efficacy against severe disease, BPBV efficacy against infection and the assumed size of the BPBV stockpile was carried out. Bars plot the mean % reduction in global COVID-19 deaths across 100 simulations, each utilising a single draw from the previously estimated posterior distribution of Rt for each country; error bars represent the 95% confidence interval for those 100 simulations. Bars are coloured according to the assumed size of the stockpile, with the x-axis varying the readiness scenario considered, facet rows varying the assumed efficacy against infection (either 0% or 35%) and facet columns the assumed efficacy against severe disease (either 50%, 70% or 95%).

**Supplementary figure 5:**
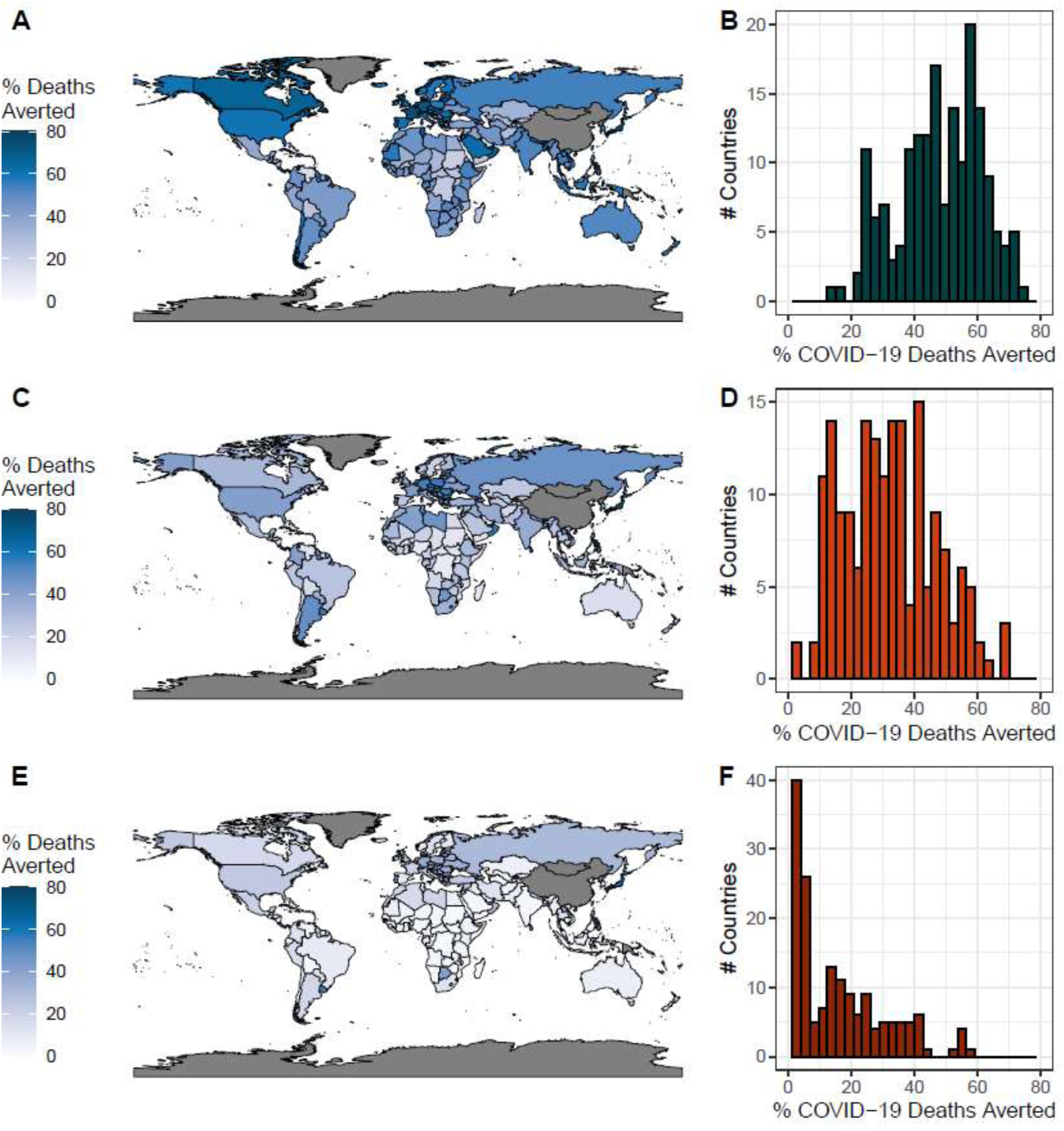
Global BPBV impact under different readiness scenarios. A) Modelled impact of the BPBV during the first year of the COVID-19 pandemic in different countries around the world, assuming the immediate readiness scenario. Colour indicates the % of COVID-19 deaths in first year of pandemic averted, if BPBV were available. B) Histogram of percentage of COVID-19 deaths averted by the BPBV in each country, for the immediate readiness scenario. C) As for (A), but assuming the Phase 3 readiness scenario. D) As for (B), but assuming the Phase 3 readiness scenario. E) As for (A), but assuming the Phase 1 readiness scenario. F) As for (B), but assuming the Phase 1 readiness scenario.

**Supplementary Figure 6.**
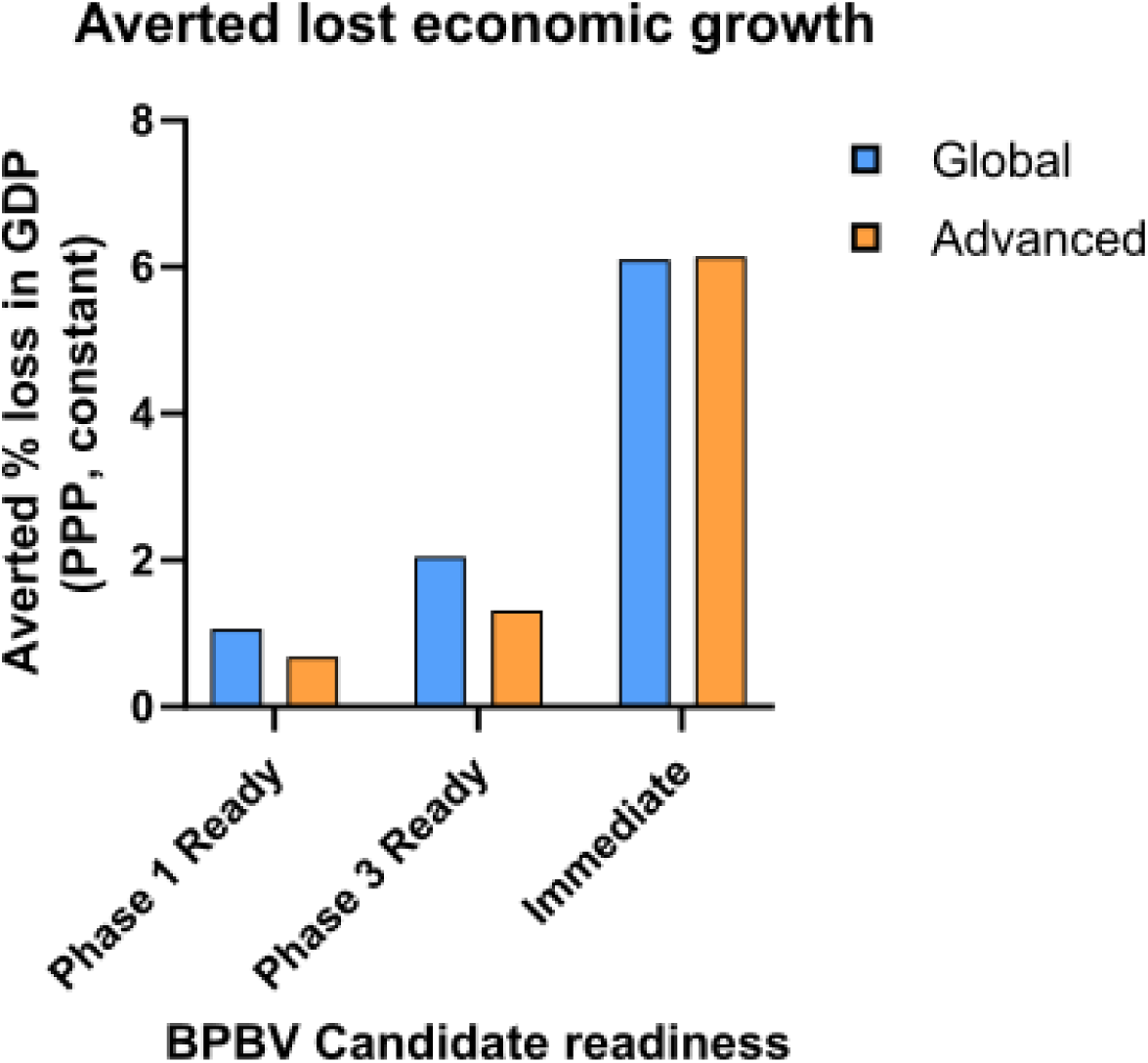
Potential BPBV impact on GDP during the COVID-19 pandemic. Immediate readiness = GDP (PPP, constant prices) grows at 2015-2019 average; Phase 3 ready = 2020 GDP change + 2.5 quarters of average growth; Phase 1 ready = 2020 GDP change + 1.3 quarters of average growth. Averted loss in global GDP indicates the relative difference between the estimated GDP growth of each scenario and the observed GDP growth during 2020. Modelling for global (blue) and advanced economies only (orange) displayed.

## Supplementary materials

### Methods

#### In silico design

Design of the S_T2_20 and S_T2_35 antigens is detailed in the cited literature ^15,16^. S_T2_37 consists of the codon-optimised sequence from MERS-CoV (Hu/Jeddah-KSA-173RS1101/2017, Accession id: QGW51898) with two mutations at Site 1060-1061 to stabilise the full-length spike in pre-fusion conformation. The sequence was further codon optimised for improved expression. The N_T2_2 antigen was designed using a phylogenetic approach. Nucleotide sequences of the nucleoprotein gene from SARS-CoV-2 and SARS-CoV were downloaded from the GISAID database. The sequences were then trimmed to the coding region and were filtered for redundancy at 95% nucleotide sequence identity. Multiple sequence alignments of NP were generated using the MAFFT algorithm with default parameters. A phylogenetic tree was produced using the previously generated multiple sequence alignment (MSA) as an input to the IQ-tree algorithm. The optimal nucleotide model for phylogenetic tree generation was chosen according to Bayesian information criteria (BIC) score. The phylogenetic tree and MSA were used as input to Hyphy (48) to generate the antigen N_T2_2 antigen. Utilizing the dataset of sequences, a unique antigen sequence was generated that was phylogenetically closest to all the input sequences in comparison to any sequence in the downloaded dataset.

#### Cells and viruses

HEK293T/17 cells were obtained from ATCC, Vero+TMPRSS2+huACE2 (VAT) were obtained from Suzannah Rihn, University of Cambridge. B.1.617.2 (POR2) was obtained from UKHSA, Porton Down. VeroE6/TMPRSS2 cells were obtained from BPS Bioscience (cat# 78081, San Diego, California, USA), and used for titration of animal samples from mouse adapted SARS-CoV and MERS-CoV (maSARS-CoV, maMERS-CoV) infections. VeroE6/TMPRSS2 cells were maintained in MEM medium supplemented with 10% fetal bovine serum (FBS), 1X MEM Non-essential Amino Acids, 1% Sodium Pyruvate, and 1% Penicillin/Streptomycin, and 3 μg/ml of Puromycin. Prior to virus titrations, growth media was removed, and replaced with 1X MEM, 1% FBS. BHK-T7 (RRID: CVCL_0059) were used for transfections in the reverse genetics construction of maSARS-CoV and maMERS-CoV and maintained in MEM supplemented with 5% FBS, 4 mmol L-Glutamine, and 1% Penicillin/Streptomycin. Vero cells (ATCC CCL-81) and VeroE6 (ATCC CRL-1586) were grown and maintained on MEM supplemented with 5% FBS, 4 mmol L-Glutamine, and 1% Penicillin/Streptomycin.

#### SARS-CoV

Mouse adapted SARS-CoV (maSARS-CoV) was made by introducing 6 nucleotide mutations into our reverse genetics system for SARS-CoV strain Tor2 (Accession AY274119). These mutations were previously identified based on mouse passaging of the Urbani strain of SARS-CoV and were shown to be lethal in BALB/c mice^32^. BHK cells were transfected with maSARS-CoV plasmid and supernatant was blind passaged onto VeroE6 cells where CPE was evident at 3 days post infection. Viral stocks of maSARS-CoV were propagated on VeroE6 cells for 3 days, clarified by centrifugation, and stored at - 80°C. A vial of frozen stock virus was thawed and titrated on Vero cells to determine the plaque forming units per mL (PFU/mL) and median tissue culture infectious dose (TCID_50_).

#### MERS-CoV

Mouse adapted MERS-CoV (maMERS-CoV) was generated using reverse genetics techniques by inserting 12 mutations into the MERS-CoV genome that had been previously identified^33^. BHK cells were transfected with maMERS-CoV plasmid and supernatant was blind passaged onto Vero cells where CPE was evident at 3 days post infection. Viral stocks of maMERS-CoV were propagated on Vero cells for 3 days, clarified by centrifugation, and stored at -80°C. A vial of frozen stock virus was thawed and titrated on Vero cells to determine the plaque forming units per mL (PFU/mL) and median tissue culture infectious dose (TCID_50_).

#### SARS-CoV-2 Delta

Delta clinical isolate (B.1.617.2, POR2) was propagated in VAT cells and quantified by standard semisolid agarose plaque assay. Briefly VAT cells were grown in DMEM/F12 supplemented with 10% FBS (12484028, Thermo Fisher) and 10,000 U/ml Penicillin/Streptomycin (15140163, Thermo Fisher) and seeded into 6-well plates to reach 70% confluence the day after seeding. Culture plates were brought into the containment level 3 laboratory and infected with a 10fold dilution series of virus for 30 minutes at room temperature with rocking. Pre-warmed semisolid agarose (16500500, Thermo Fisher) heated to 50°C was then added at 2% (final concentration) in DMEM/F12 directly onto wells and incubated for 48h. 10% neutral buffered formalin (HT501128-4L, Merck) was then added to inactivate virus and fix cells for 1h at room temperature. Liquid was discarded and 0.25% crystal violet solution (in 20% ethanol) added to wells and stained for 30 minutes. Liquid was then removed before plates were plunged twice into 1% sodium hypochlorite and then sterile water. Viral titres were calculated as PFU/ml, and downstream mice were infected with 1x10^4^ PFU in a volume of 20µl. All viruses were sequenced by next generation sequencing^34^ following propagation and no mutations were identified in the stocks compared to the parental sequences.

#### Virus production and titration info, dosing information for mice (SARS-CoV and MERS-CoV)

For maSARS-CoV challenges, BALB/c mice were anesthetized with gaseous isoflurane and intranasally (IN) infected with 10 LD_50_ (3.98 x 10^5^ TCID_50_/mL) in 50μL MEM/BSA evenly divided between the nares. MERS-CoV susceptible mice were generated by introducing the human amino acid substitutions 288L and 330R in the dipeptidyl peptidase 4 (DPP4) gene of C57BL/6 mice^35^. huDPP4 mice were anesthetized as above and IN infected with 7 LD_50_ (3.16 x 10^4^ TCID_50_/mL) of maMERS-CoV in 50μL MEM/BSA evenly distributed between the nares.

#### mRNA production

Sequences for mRNA vaccine antigens (S_T2_20, S_T2_35, S_T2_37 and N_T2_2) were synthesised by *in vitro* transcription (IVT) from linearised plasmid DNA templates using modified nucleotides, generating partial modified mRNAs. IVT was carried out for 120 m at 37 °C using T7-RNA polymerase and co-transcriptional capping via Anti-Reverse Cap Analog (ARCA). This was followed by digestion of the template using DNAse I. After IVT, dephosphorylation of mRNAs was carried out for 15 min at 37 °C using alkaline phosphatase and then these were enzymatically polyadenylated for 10-30 min at 37 °C using PolyA polymerase, resulting in a Poly A tail of approximately 120 nucleotides. Purification was carried out by precipitation and then mRNAs were formulated in water at a final concentration of 1 mg/mL. mRNAs were stored at -80 °C until encapsulation. Each mRNA was encapsulated into ionisable lipidoids via nanoprecipitation using microfluidic mixing of mRNA in citrate buffer (pH 4.5) with ionisable-, structural-, helper- and polyethylene glycol (PEG) lipids in ethanol. This was followed by buffer exchange and concentration via tangential flow filtration. mRNA LNPs were 0.2μm membrane filtered and stored at -20 °C until use. After manufacturing and freezing, the final encapsulated mRNAs were analytically characterised for particle size (≤ 100 nm), particle polydispersity (≤ 0.20), encapsulation efficiency (≥ 90%), mRNA integrity (≥ 90%) and mRNA identity (confirmed length).

#### Statistical analysis

Two-tailed Mann–Whitney U tests were carried out for all neutralising antibody group comparisons using GraphPad Prism 10.4.

**Supplementary Table 1.**
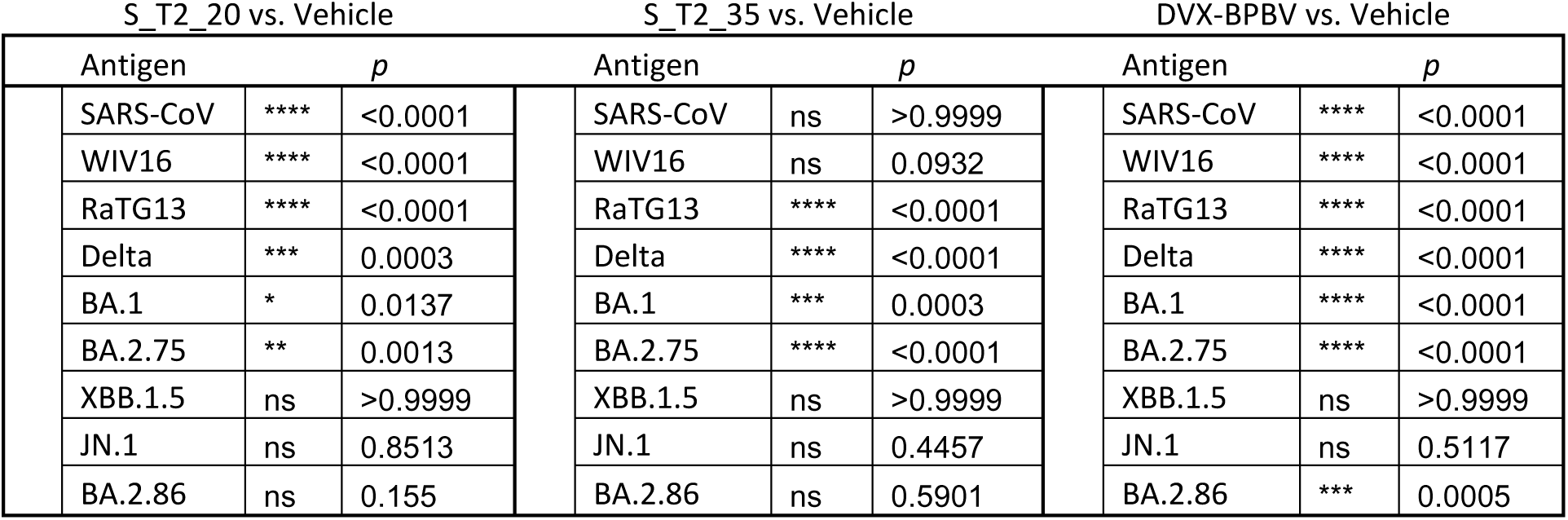
Statistical comparisons (p values) between vaccine groups displayed in Figure 2, sarbecovirus pseudotype neutralisation data. Comparisons between vaccines and the naïve (vehicle) group, as well as the DVX-BPBV against individual vaccines are displayed.

**Supplementary Table 2.**
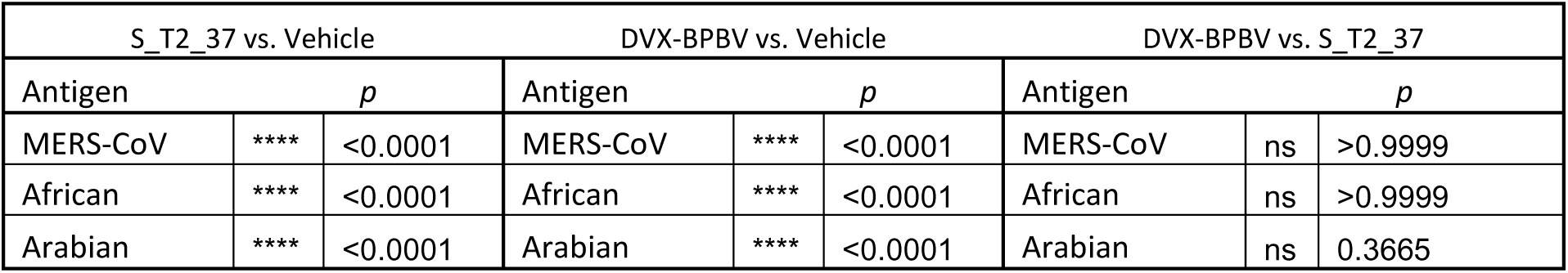
Statistical comparisons (p values) between vaccine groups displayed in Figure 2, merbecovirus pseudotype neutralisation data. Comparisons between vaccines and the naïve (vehicle) group, as well as the DVX-BPBV against individual vaccines are displayed.

**Supplementary Table 3.**
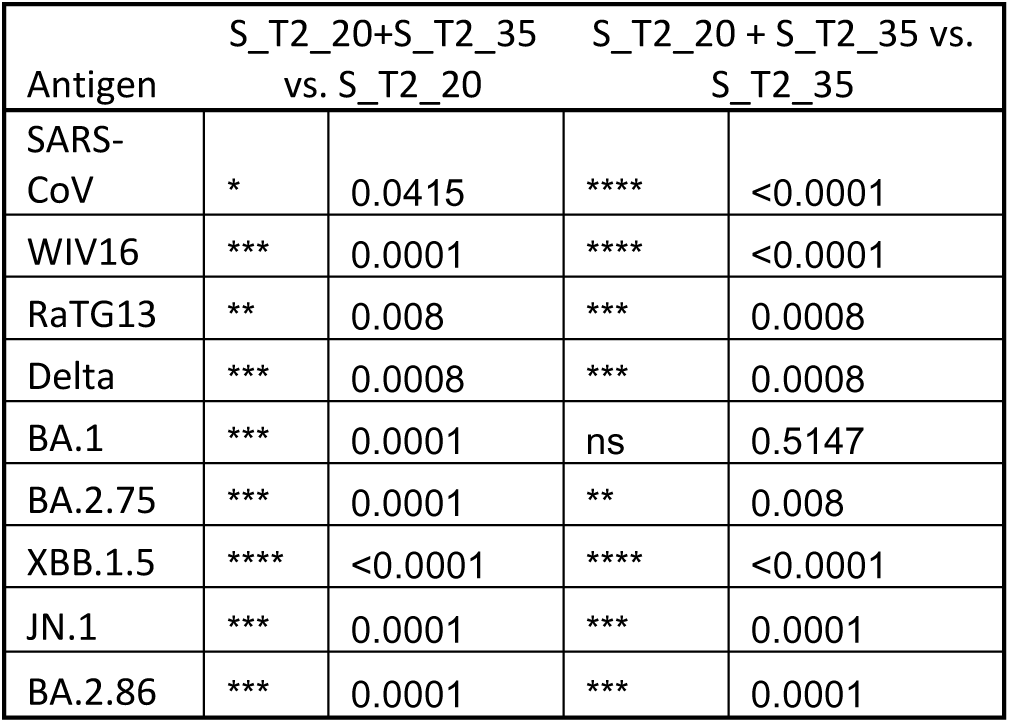
Statistical comparisons (p values) between vaccine groups displayed in Figure 2B and 2D, sarbecovirus pseudotype neutralisation data. Comparisons between Hartley guinea pigs immunised with S_T2_20 and S_T2_35, and mice immunised with S_T2_20 or S_T2_35 alone.

#### Animal work (BALB/c and C57BL/6 Mice, Hartley Guinea Pigs)

Immunogenicity animal work was conducted at University Biomedical Services, Cambridge, U.K, under approved project licence P8143424B and with ethical clearance from the Animal Welfare and Ethical Review Board of the University of Cambridge. Challenge studies for SARS-CoV-2 Delta (B.1.617.2) were also carried out in Cambridge. Challenge studies for the SARS-CoV and MERS-CoV arms of the preclinical efficacy studies were carried out at the National Microbiology Laboratory (NML), Winnipeg, Canada with approval from the Animal Care Committee (ACC) at the Canadian Science Centre for Human and Animal Health (CSCHAH) under approved Animal Use Document H-20-011 (SARS-CoV) and H-22-005 (MERS-CoV). All infectious work with maSARS-CoV and maMERS-CoV was performed in the containment level 4 (CL-4) laboratory at the NML. Animals were acclimatized for 5-7 days before beginning the experiments and were provided food and water *ad libitum* throughout the experiment. All animals were monitored and weighed daily throughout the experiments and scored for signs of disease.

Animal immunogenicity experiments were carried out in 7-week-old female BALB/c mice (Charles River Laboratories, U.K). Briefly two doses of mRNA vaccine were administered with a three-week interval. Blood was taken through the saphenous vein route at three weeks after first dose as well as three and six weeks after second dose, the latter being obtained by cardiac puncture under non-recovery anaesthesia. Confirmatory and synergy experiments were carried out in outbred female 7-week-old Hartley guinea pigs (Inotiv CRO, Netherlands). Briefly, guinea pigs were immunised twice with mRNA vaccines with a three-week interval. Blood was taken six weeks after boost by cardiac puncture under non-recovery anaesthesia. For preclinical efficacy studies, K18-huACE2 (JAX via Charles River Laboratories, Paris) were used for SARS-CoV-2 Delta (B.1.617.2) at University Biomedical Services, Cambridge, United Kingdom. BALB/c mice (Charles River) were used for all maSARS-CoV infections, and huDPP4 transgenic mice (University of Manitoba) were used for maMERS-CoV challenges. All studies followed the same schedule with two doses of mRNA vaccine with a three-week interval. Mice were sampled at three weeks post prime and three weeks post boost and then challenged with a lethal dose four weeks after boost. The challenge period lasted 14 days and half of each respective group (6/12) were culled on day three to measure viral titres in the lungs.

**Supplementary Table 4.**
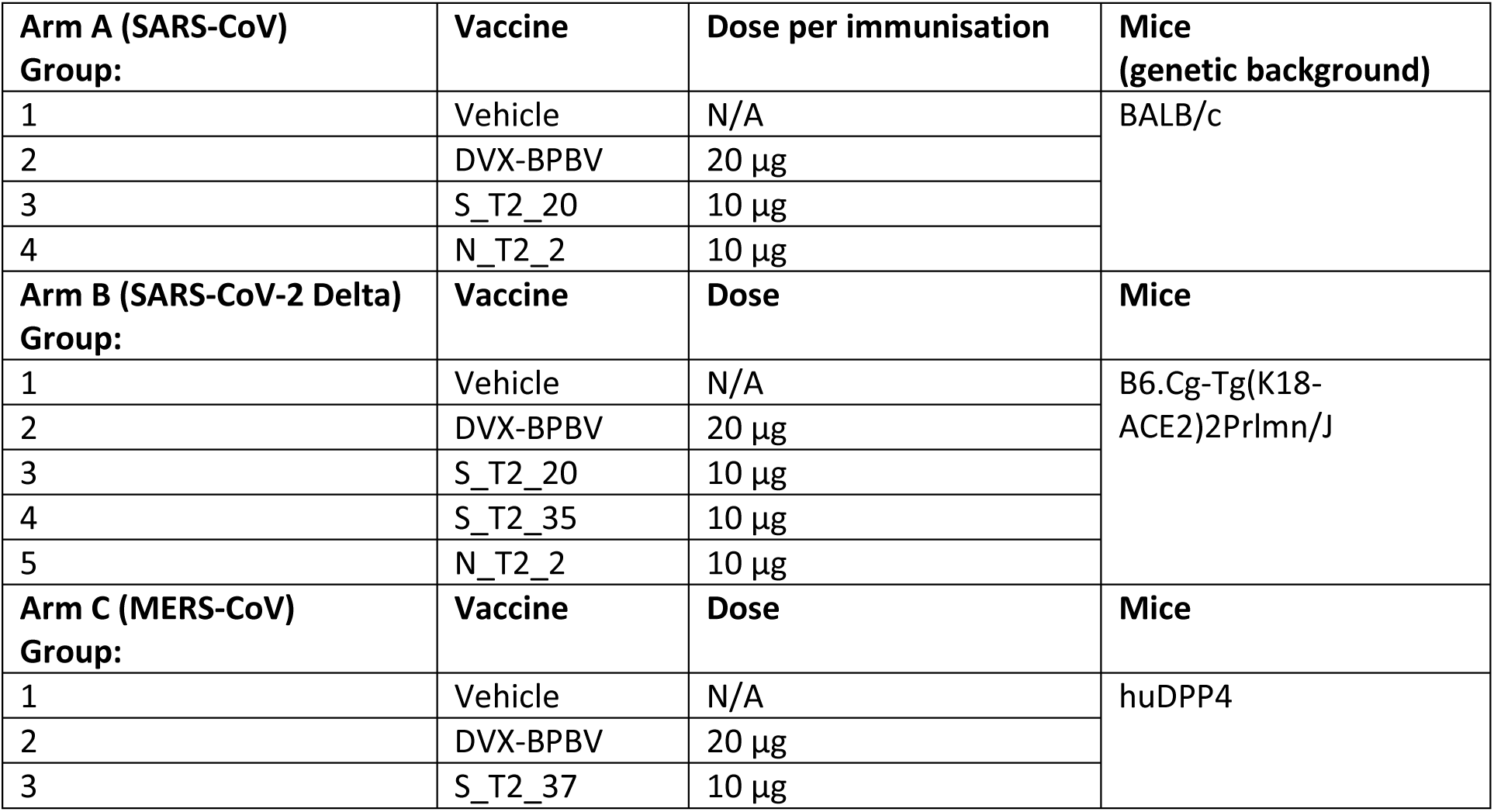
Group organisation (n=12 per group) for vaccinated (2 doses total with 3-week interval) mice challenged with SARS-CoV (A), SARS-CoV-2 Delta (B) and MERS-CoV (C). Mice receiving single antigen doses received 10µg total mRNA, while DVX-BPBV immunised mice received 20µg total mRNA (5µg per antigen).

**Supplementary Table 5.**
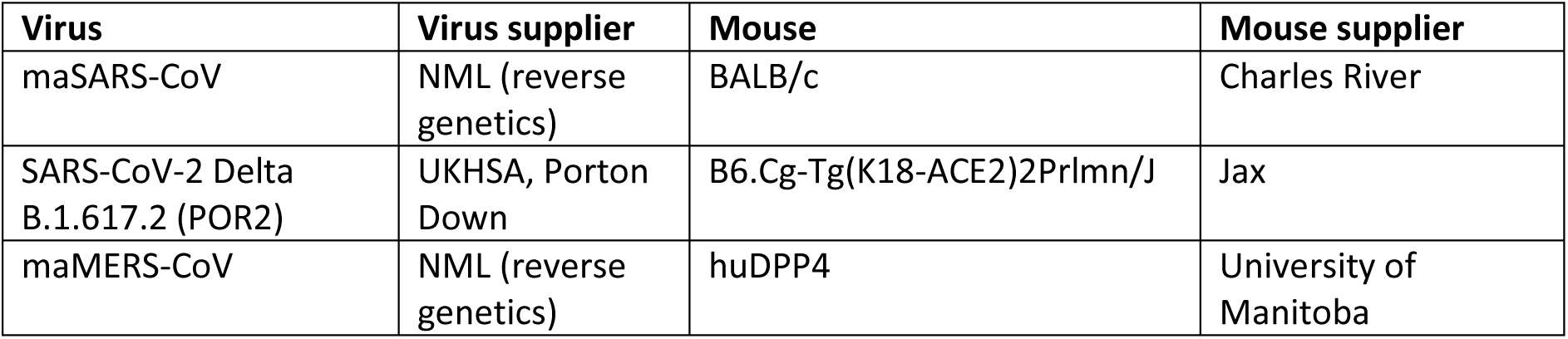
Viruses, mouse strains and suppliers used in animal infection studies.

#### SARS-CoV-2 TCID50

TCID_50_ values were generated from homogenised mouse lungs for all samples. 96-well cell culture plates were used to culture VAT cells, and eight tenfold dilutions of lung homogenate (diluted in DMEM with 2% FCS) were titrated (1:10 start) on the cells in triplicate. At 48h cells were checked for CPE and then fixed with 10% neutral buffered formalin, stained with crystal violet and left to dry overnight. Wells were scored for CPE using a discontinuous, binary scoring system and a modified Reed and Muench method^36^ was used to determine TCID_50_.

#### SARS-CoV and MERS-CoV TCID50

A section of mouse lung tissue from necropsies conducted on day 3 post maSARS-CoV and maMERS-CoV infection were weighed and placed in 1 mL of MEM with 1% Penicillin/Streptomycin. A 5 mm stainless steel bead was added and samples were homogenized using the Bead Ruptor Elite Tissue Homogenizer (Omni) for 30 seconds at 4 m/s. Homogenates were clarified by centrifugation at 1500 x g for 10 minutes, and 10-fold serial dilutions were made in MEM supplemented with 1% FBS and 2% Penicillin/Streptomycin. Dilutions were added in triplicate to 96 well plates seeded with VeroE6/TMPRSS2 cells (BPS Bioscience) and incubated at 37°C, 5% CO_2_. At 4 days post infection, the cells were examined for CPE, and TCID_50_ titers were calculated as described above.

#### Pseudotyped lentivirus production

Spike bearing lentiviral pseudotypes were produced by transient transfection of HEK293T/17 cells with packaging plasmids p8.91 and pCSFLW and different spike-bearing expression plasmids in the pEVAC plasmid backbone, using the Fugene-HD (Promega, E2311) transfection reagent. Supernatants were harvested after 48 h, passed through a 0.45 µm cellulose acetate filter, and titrated on HEK293T/17 cells transiently expressing either human ACE-2 or DPP4, and human TMPRSS2. Target HEK293T/17 cells were transfected 24 h prior to infection with 2 µg pCAGGS-huACE-2 or pCAGGS-huDPP4 and 150 ng pCAGGS-TMPRSS2 in a T75 tissue culture flask.

#### Pseudotype based microneutralisation assay

Pseudotype-based micro-neutralisation assays (pMN) were performed as described previously^18^. Serial dilutions of serum were incubated with lentiviral pseudotypes bearing sarbecovirus or merbecovirus spike proteins for 1 h at 37 °C, 5% CO_2_ in 96-well white cell culture plates. 1.5 × 10^4^ HEK293T/17 cells transiently expressing human ACE-2 or DPP4 and TMPRSS2 were then added per well and plates incubated for 48 h at 37 °C, 5% CO_2_ in a humidified incubator. Bright-Glo (Promega) was then added to each well and luminescence read after a five-minute incubation period. Experimental data points were normalised to 100% and 0% neutralisation using cell only and virus positive cell control wells respectively and non-linear regression analysis performed in GraphPad Prism 10 to produce neutralisation curves and resulting IC_50_ values. Antisera generated from a pool of XBB.1.5 spike mRNA immunised guinea pigs were used as an internal control for inter plate and virus variation.

#### T-cell assays

##### Flow cytometry cytokine assay

Under sterile conditions murine splenocytes were placed into RPMI (10% FBS, 1% Penicillin/Streptomycin 50 µM beta-mercaptoethanol) media in a 96 well U-bottomed plate at a concentration of 2.25x10^6^/ml, 200 µl per well. The cells were incubated either with Protein Transport Inhibitor (ThermoFisher 00-4980-03) + 1 µg/ml SARS-CoV Spike or NP, SARS-CoV-2 Spike or NP, or MERS-CoV Spike pooled peptide. PTI and CSC were used as a negative and positive control respectively. Cells were incubated at 37°C for 6 hours.

Cells were washed with FACS buffer (PBS, 0.5% BSA, 0.01% NaN_3_) three times, blocked with FACS buffer with 2% heat inactivated rat serum for 15 minutes at 4°C, then stained with Fixable Viability Dye eFluor™ 780 (ThermoFisher 65-0865-14), CD4 Monoclonal Antibody (RM4-5) APC (ThermoFisher 17-0042-83), CD3e Monoclonal Antibody (145-2C11) PE-Cyanine5.5 (ThermoFisher 15-0031-82) and CD8a Monoclonal Antibody (53-6.7) PE-Cyanine7 (ThermoFisher 25-0081-82). Cells were incubated for 30 minutes at 4°C in the dark, washed three times with FACS buffer, then resuspended in CytoFix/CytoPerm (BD Biosciences 554714) and left at 4°C in the dark for 20 minutes. Cells were then washed with permeabilisation/wash buffer 3 times, stained for intracellular cytokines with IFNγ monoclonal antibody (XMG1.2) Alexa Fluor™ 488 (ThermoFisher 53-7311-82) and TNFα monoclonal antibody (MP6-XT22) PE-eFluor™ 610 (ThermoFisher 61-7321-82) at room temperature in the dark for 30 minutes. Washed with permeabilisation/wash buffer 3 times and finally resuspended in FACS buffer for analysis on a BD LSRFortessa^TM^ HTS Cell Analyser with 5 lasers (BD Biosciences). UltraComp eBeads™ Compensation Beads (ThermoFisher 01-2222-42) were used for single colour compensation controls. Analysis was performed using FlowJo 10.9.0 (BD Biosciences).

##### Peptide design and synthesis

Peptide sequence predictions for wild type proteins were generated using NetMHCpan and NetMHCIIpan software without a length restriction setting (https://services.healthtech.dtu.dk/services/NetMHCIIpan-4.3/^37,38^). For predicted peptide pools, the library of peptides was filtered for binding values, number of alleles being targeted, and the number of times amino acid was predicted to be a part of the peptide. Once such assembly of peptides was selected, the predicted peptides were aligned to the reference sequence and a peptide library of 15mer with overlap of 11mer was generated. For overlapping peptide pools, entire length of the protein reference sequence was divided into 15mer peptides with 11mer overlap. All peptides were synthesized by GenScript and reconstituted in 4% DMSO in PBS.

##### Isolation of murine splenocytes

Murine splenocytes were isolated from dissected spleens by mashing spleens on MACS SmartStrainers (Miltenyi Biotec) and layering them on the Histopaque 1083 reagent (SigmaAldrich). buffy coat containing immune cells was collected from the phase separation region and washed in R10 medium (RPMI medium supplemented with 10% FBS). Cell pellets were resuspended and frozen in FBS supplemented with 10% DMSO Hybri-Max (SigmaAldrich).

##### ELISpot analysis of murine splenocytes

ELISpot assays with cryopreserved splenocytes were performed according to the manufacturer’s protocol using the mouse IFN-γ Single-Color ELISpot kit (CTL™ ImmunoSpot). Splenocytes were thawed, washed once with R10 medium and resuspended in CTL™ medium supplemented with L-glutamine at the desired cell density (in the range of 2-5x10^6^ cells/mL). Custom made overlapping peptides for SARS-CoV-2 NP (NCBI reference sequence: YP_009724397.2) and SARS-CoV NP (NCBI Reference Sequence: YP_009825051.1) were used. For the stimulation, 100 µL of peptide solution in CTL at 2 µg/peptide/mL were mixed with 100 µL of cell suspension in the 96-well ELISpot plate and incubated for 24h in 37°C in humidified incubator with 5% CO_2_. As a positive control, eBioscience cell stimulation cocktail (ThermoFisher Scientific) was used. CTL medium was used as a non-stimulation control. The plates were developed as per manufacturer’s protocol, dried, scanned and analysed on ELISpot reader S6 Ultra M2 (ImmunoSpot).

##### Phylogenetic analysis

Amino acid sequences from the spike proteins for SARS-CoV-1, WIV16, RaTG13, Delta, BA.1, BA.2.75, XBB1.5, JN.1, BA.2.86, MERS-CoV, D998/15 and 011/DAB/C8/F<1 were downloaded from Genbank, and a maximum likelihood tree was constructed using MAFFT v7 with auto algorithm to align sequences^39^. Gaps in alignments were trimmed using TrimAl^40^. The resulting tree was built using IQ-TREE2 with automatic model selection with 1000 ultrafast bootstrap replicated to assess branch support^41^.

##### Summary of Modelling Framework

We explored the potential utility of the BPBV in vaccination campaigns focussed on rapid mass-vaccination of priority groups following pathogen detection to support disease burden reduction and relaxation of societal restrictions imposed to control transmission (as has been the case with SARS-CoV-2 vaccination campaigns). To do this, we adapted a previously published dynamical model of SARS-CoV-2 transmission used to evaluate and explore the impact of SARS-CoV-2 transmission on COVID-19 mortality during the pandemic^22,42^, and which has more recently been used to explore a range of different implementation strategies for a BPBV^43^. Briefly, the model is an age-stratified SEIRS (susceptible-exposed-infectious-recovered-susceptible) model that explicitly models the progression of COVID-19 disease severity, the transition through various levels of healthcare, and the introduction of vaccination campaigns. Complete details of the previous versions of the model are given in ^14,15,16,17^.

##### Retrospective Evaluation of Potential Impact during SARS-CoV-2 Pandemic

Using previously published model fits calibrated within a Bayesian framework to excess mortality data (known to be a more complete measure of pandemic mortality, especially in LMIC settings with less robust vital registration)^21,45^, we explored the potential impact that a stockpiled BPBV could have had on COVID-19 mortality in the first year of the pandemic. We used the resulting model fits to estimate the time-varying reproductive number, Rt, and its associated uncertainty by sampling 100 draws from the estimated posterior distribution of Rt from these previous fits. To estimate the impact of BPBV, we simulated a counterfactual scenario for each sampled Rt trajectory in which the BPBV was introduced after some delay following the 100^th^ globally reported COVID-19 death. We varied the length of this delay to reflect 3 different scenarios reflecting different levels of pre-pandemic readiness:

- “High Readiness”: The BPBV is stockpiled, has been clinically evaluated and is available for immediate administration to eligible individuals following stockpile activation (i.e. after the 100th globally reported COVID-19 death).
- “Moderate Readiness”: The BPBV is stockpiled and has been clinically evaluated to the point where Phase 3 trials can immediately be initiated following stockpile activation (i.e. after the 100th globally reported COVID-19 death).
- “Low Readiness”: The BPBV has been evaluated to only a minimal degree, and Phase 1, 2 and 3 trials are required following stockpile activation before it can be administered to eligible individuals.

In addition to this, we also varied the size of the assumed country-specific BPBV stockpile: assuming either that i) all countries have access to a BPBV stockpile sufficient to vaccinate 50% of their eligible elderly population at a rate representing the average COVID-19 vaccination rate for each World Bank Income Strata based on data from Our World In Data ^23^; ii) all countries have access to a BPBV stockpile sufficient to vaccinate 90% of their eligible elderly population at a rate representing the average COVID-19 vaccination rate for each World Bank Income Strata based on data from Our World In Data ^23^; or iii) countries have access to a BPBV stockpile sufficient to vaccinate 20%/40%/60% or 80% of their eligible elderly population at a rate representing the average COVID-19 vaccination rate for each World Bank Income Strata based on data from Our World In Data^23^, with the size of the stockpile depending on World Bank Income Strata (LIC, LMIC, UMIC and HIC respectively). We also varied assumptions about BPBV efficacy against severe disease (50%, 75% or 95%) and infection (0% or 35%). All of these analyses were carried out under the strong assumption that availability of the BPBV would not have altered the NPIs imposed in response to SARS-CoV-2 (and hence alterations to Rt due to NPIs would be the same across both scenarios). We then calculated the deaths averted as a result of BPBV vaccination by subtracting the estimated COVID-19 deaths from the simulation with BPBV vaccines included from the estimated COVID-19 deaths from the simulation with only the real-world vaccination campaign included and reported the median deaths averted per 1,000 population.

**Supplementary table 6:**
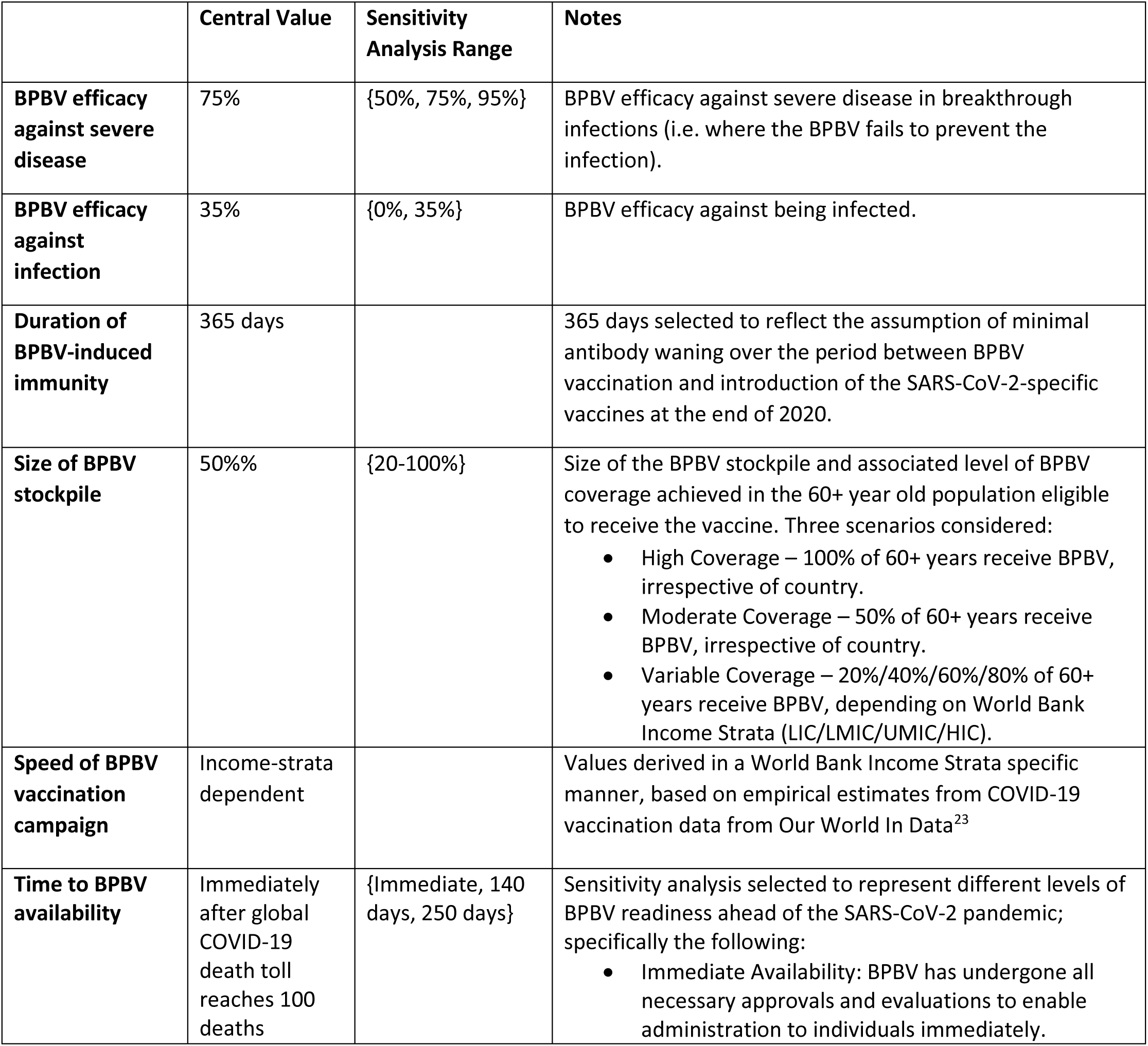
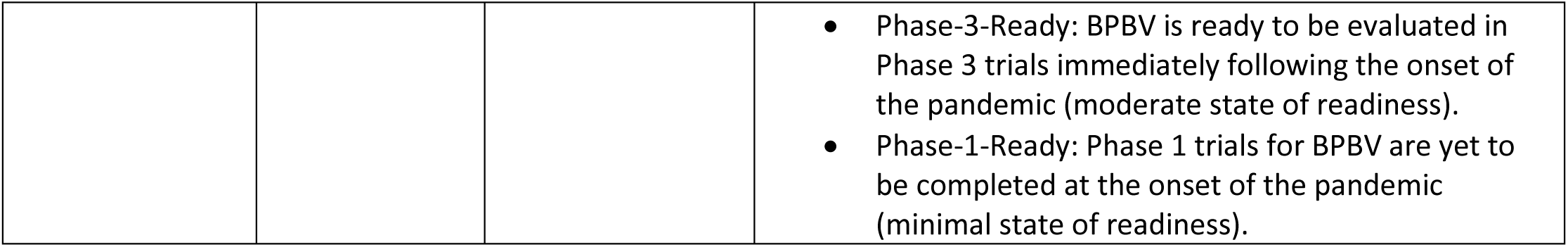
Description of model parameters varied in dynamical compartmental modelling of mass-vaccination of high-risk populations with BPBV.

##### Summary of economical modelling (Airfinity)

The 5-year pre-COVID-19 pandemic average (2015-2019) percentage growth of real (constant prices) GDP at purchasing power parity (PPP) per annum and per quarter were calculated using data obtained from the IMF. GDP change due to the COVID-19 pandemic was defined as the observed real GDP (PPP) change in 2020. For expected GDP change under the “immediate readiness” scenario, the average pre-COVID-19 annual growth was applied. For the other scenarios expected growth was calculated as GDP change during the pandemic plus normal growth applied on a pro-rata basis of how long left in a year from time of anticipated vaccine rollout - 225 days (2.5 quarters) for “Phase 3 ready” and 115 days (1.3 quarters). Averted % loss in GDP was calculated as the difference between observed 2020 growth and expected growth from each scenario.

## Notes

### Author Declarations

We used only openly available, published human data: Global excess deaths associated with COVID-19 (modelled estimates). Accessed July 21, 2025. https://www.who.int/data/sets/global-excess-deaths-associated-with-covid-19-modelled-estimates Global excess deaths associated with COVID-19 (modelled estimates). Accessed July 21, 2025. https://www.who.int/data/sets/global-excess-deaths-associated-with-covid-19-modelled-estimates

