## Supplementary and methods for "Efficacy and potential human and economic impact of a broadly protective betacoronavirus vaccine"

One Sentence Summary: Broadly protective betacoronavirus vaccine protects mice from lethal challenge by SARS-CoV, SARS-CoV-2 and MERS-CoV, and epidemiological modelling reveals the potential human and economic impact by such a vaccine linked to its clinical readiness.

George W. Carnell[1,2,3], Robert Vendramelli[4], Charles Whittaker[5], Jonathan A. Holbrook[2], Srivatsan Parthasarathy[2], Hazel Stewart[6], Joey Olivier[1], Bryce Warner[4], Thang Truong[4], Charlotte George[1], Sneha B. Sujit[2], Sruthika K. Ashokan[2], Maria Suau Sans[1], Chloe Qingzhou Huang[1], David A. Wells[2], Paul Tonks[1], Martina Pfranger[7], Sebastian Einhauser[7], Patrick Neckermann[7], Diego Cantoni[8], Andrew C. Y. Chan[1], Laura O'Reilly[1], Luis Ohlendorf[1], Isaac Unwins[9], Stefan P. Rautenbach[9], Matteo Ferrari[2], Andrew E. Firth[6], Johannes Geiger[10], Christian Dohmen[10], Verena Mummert[10], Anne Rosalind Samuel[10], Christian Plank[10], Joanne Marie M. Del Rosario[2], Nigel Temperton[11], Benedikt Asbach[7], Simon D. W. Frost[2], Rebecca Kinsley[2], Sneha Vishwanath[1,2], Sofiya Fedosyuk[2], Ralf Wagner[2,7,12], Darwyn Kobasa[4], Jonathan L. Heeney[1,2*].

1. Lab of Viral Zoonotics, Department of Veterinary Medicine, University of Cambridge, Cambridge, UK
2. DIOSynVax Ltd, University of Cambridge, Cambridge, UK
3. One Virology, Wolfson Centre for Global Virus Research, School of Veterinary Medicine and Science, University of Nottingham, Nottingham, UK
4. Special Pathogens Program, National Microbiology Laboratory, Public Health Agency of Canada, Winnipeg, Canada
5. MRC Centre for Global Infectious Disease Analysis, School of Public Health, Imperial College London, London, UK
6. Department of Pathology, University of Cambridge, Cambridge, UK
7. Institute of Medical Microbiology and Hygiene, University of Regensburg, Regensburg, Germany
8. MRC-University of Glasgow Centre for Virus Research, University of Glasgow, Glasgow, UK
9. Airfinity Ltd, London, UK
10. Ethris GmbH, Planegg, Germany
11. Viral Pseudotype Unit, Medway School of Pharmacy, University of Kent and University of Greenwich, Chatham, UK
12. Institute of Clinical Microbiology and Hygiene, University Hospital Regensburg, Regensburg, Germany

Supplementary Figure 1

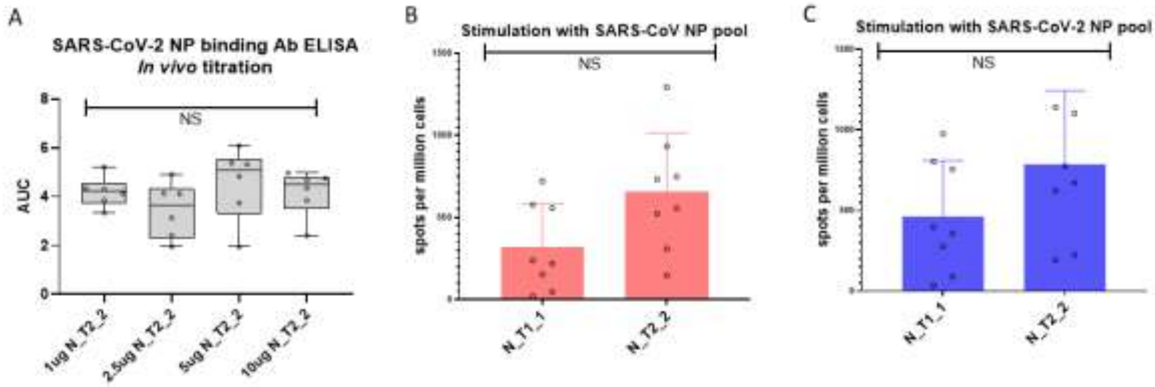


**Supplementary figure 1. Immunogenicity data for mRNA pan-sarbecovirus nucleoprotein antigen N_T2_2.**

**A)** SARS-CoV-2 Wuhan spike direct ELISA binding analysis of the serum generated by an *in vivo* titration of the N_T2_2 mRNA vaccine in BALB/c mice, with mice (n=6 per group) receiving 1, 2.5, 5 or 10µg of mRNA_N_T2_2. Area under the curve for 8-point titration of serum shown. **B)** and **(C)**, IFNγ ELISpot analysis of BALB/c mice immunised with N_T2_2 or SARS-CoV-2 nucleoprotein (N_T1_1) mRNA (10µg) stimulated with peptide pools from SARS-CoV NP (red) or SARS-CoV-2 NP (blue). All statistical comparisons using two-tailed Mann-Whitney, NS = not significant.

Supplementary figure 2.

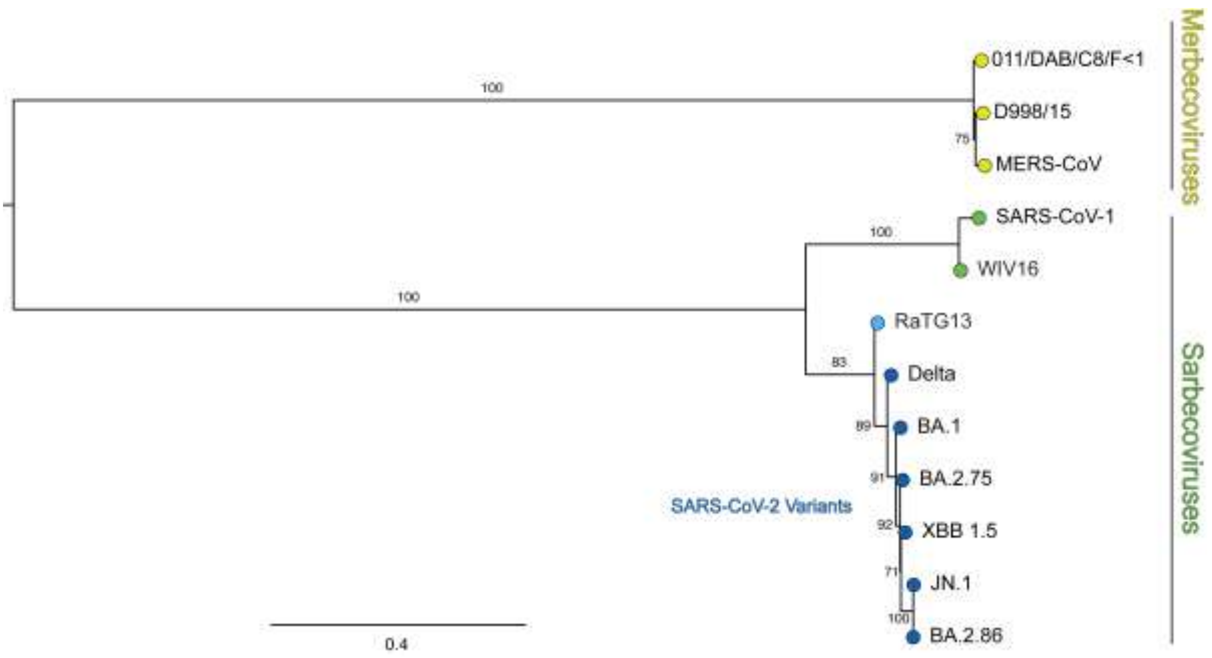


**Supplementary figure 2. Phylogeny of the Spike protein amino acid sequences used to assess the broadly protective neutralising antibodies from immunised animals**

Amino acid sequences representing the spike proteins used in this study were taken from Genbank to produce a maximum likelihood tree, using MAFFT v7 with auto algorithm to align sequences, of which gaps in the alignments were trimmed using TrimAl, and the resultant tree built using IQ-TREE2 with automatic model selection with 1000 ultrafast bootstrap replicated to assess branch support.

**Supplementary Figure 3**

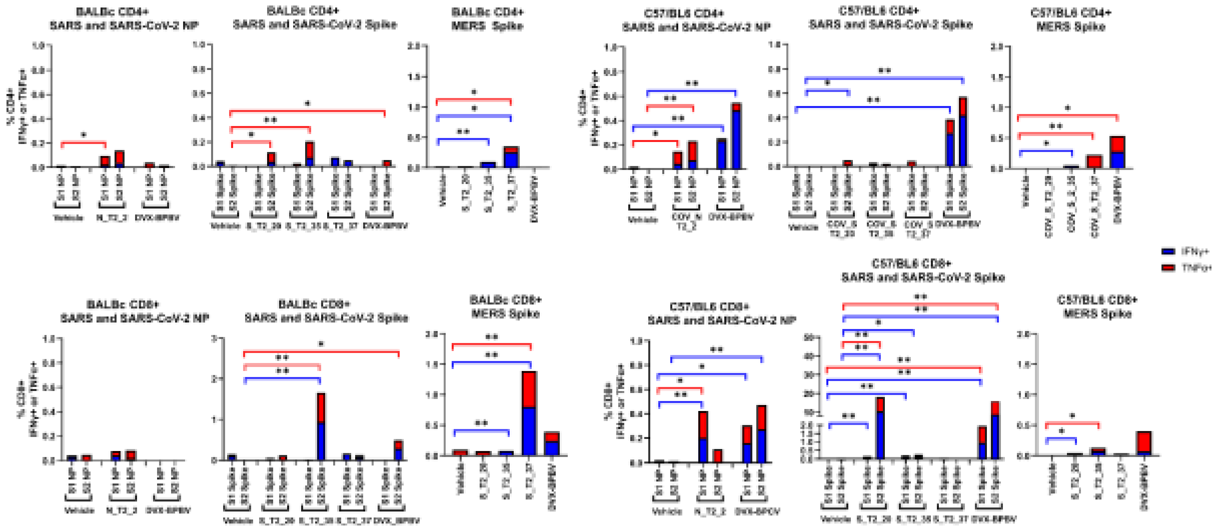


Supplementary figure 3. Flow cytometry-based T-cell analysis from BALB/c or C57BL/6 mice immunised single antigen mRNAs.

**A)** BALB/c CD4+ responses against spike or nucleoprotein peptide pools from SARS-CoV, SARS-CoV-2 or MERS-CoV. **B)** CD4+ responses from vaccinated and naïve (vehicle) immunised C57BL/6. **C)** CD8+ responses from vaccinated and naïve immunised BALB/c and **(D)** CD8+ responses from vaccinated and naïve immunised C57BL/6.

Supplementary Figure 4

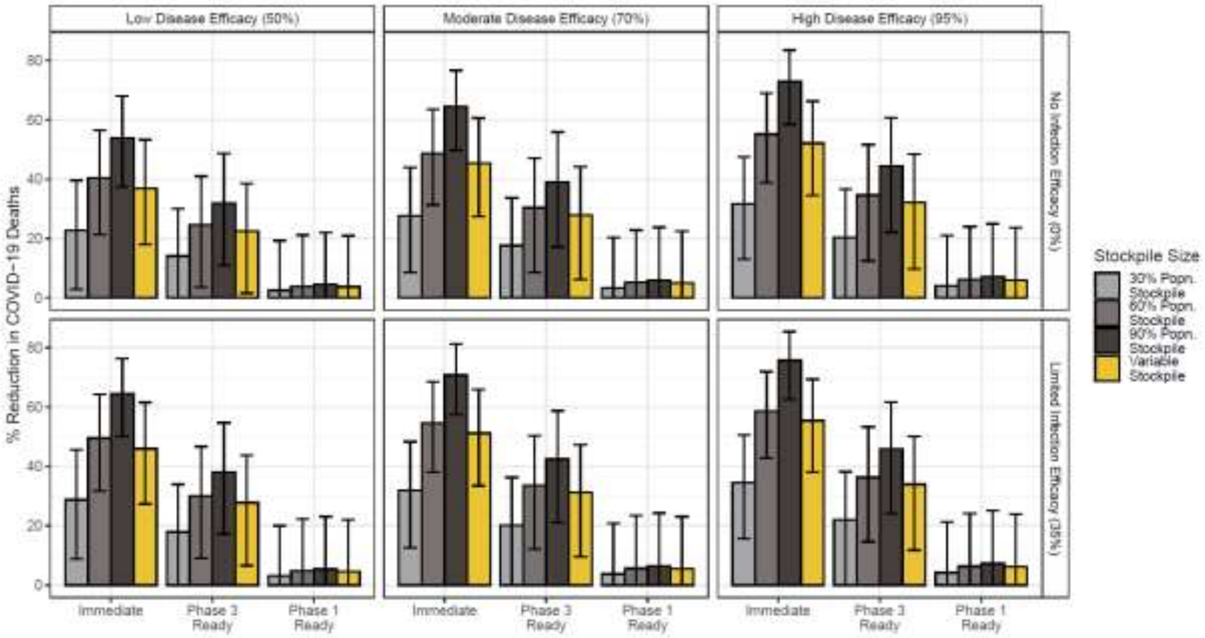


**Supplementary figure 4: Sensitivity analysis exploring the drivers of BPBV impact**

A comprehensive suite of sensitivity analyses varying readiness scenario, BPBV efficacy against severe disease, BPBV efficacy against infection and the assumed size of the BPBV stockpile was carried out. Bars plot the mean % reduction in global COVID-19 deaths across 100 simulations, each utilising a single draw from the previously estimated posterior distribution of Rt for each country; error bars represent the 95% confidence interval for those 100 simulations. Bars are coloured according to the assumed size of the stockpile, with the x-axis varying the readiness scenario considered, facet rows varying the assumed efficacy against infection (either 0% or 35%) and facet columns the assumed efficacy against severe disease (either 50%, 70% or 95%).

Supplementary Figure 5

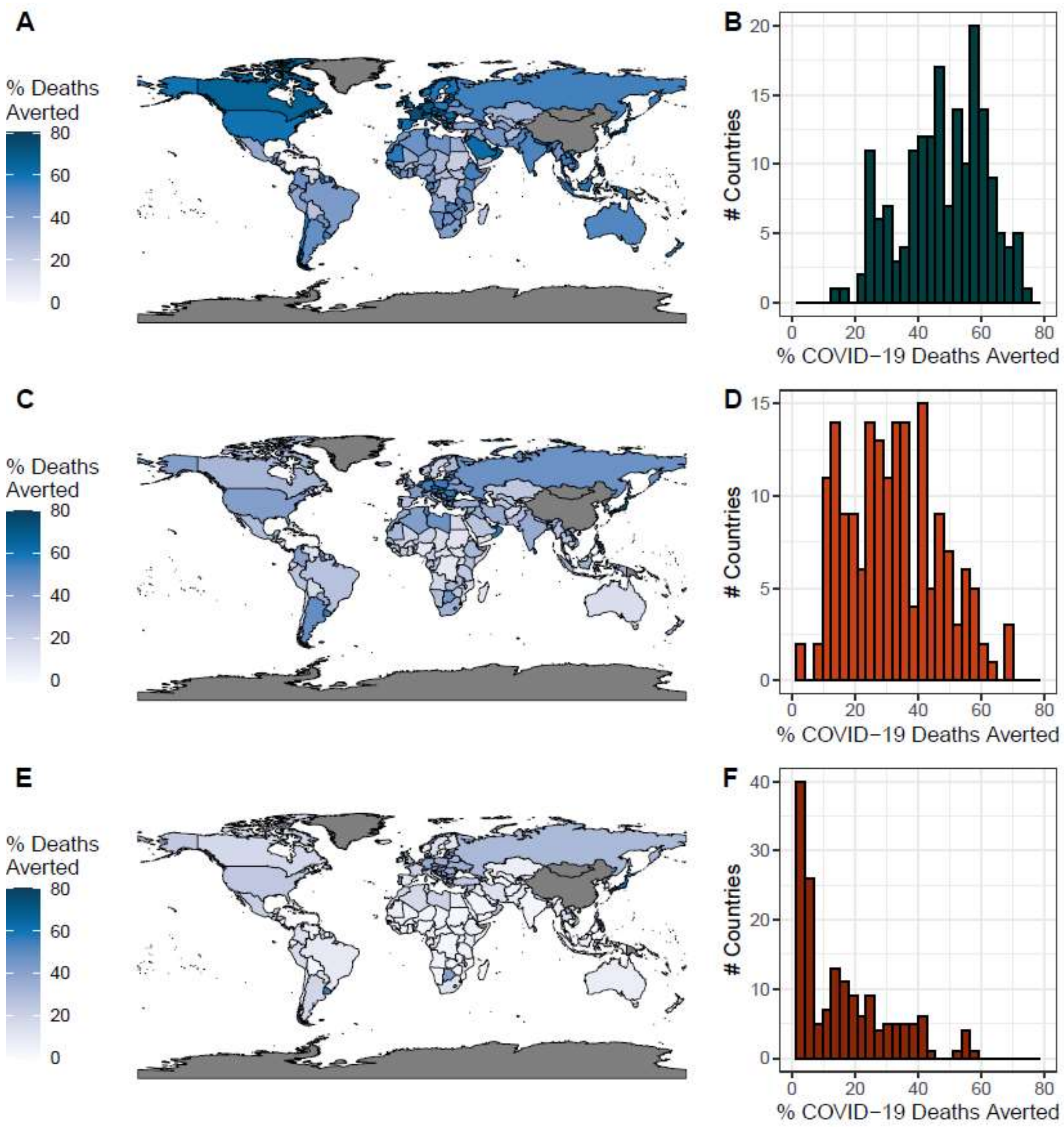


**Supplementary figure 5: Global BPBV impact under different readiness scenarios**

**A)** Modelled impact of the BPBV during the first year of the COVID-19 pandemic in different countries around the world, assuming the immediate readiness scenario. Colour indicates the % of COVID-19 deaths in first year of pandemic averted, if BPBV were available. **B)** Histogram of percentage of COVID-19 deaths averted by the BPBV in each country, for the immediate readiness scenario. **C)** As for **(A)**, but assuming the Phase 3 readiness scenario. **D)** As for **(B)**, but assuming the Phase 3 readiness scenario. **E)** As for **(A)**, but assuming the Phase 1 readiness scenario. **F)** As for **(B)**, but assuming the Phase 1 readiness scenario.

Supplementary Figure 6.

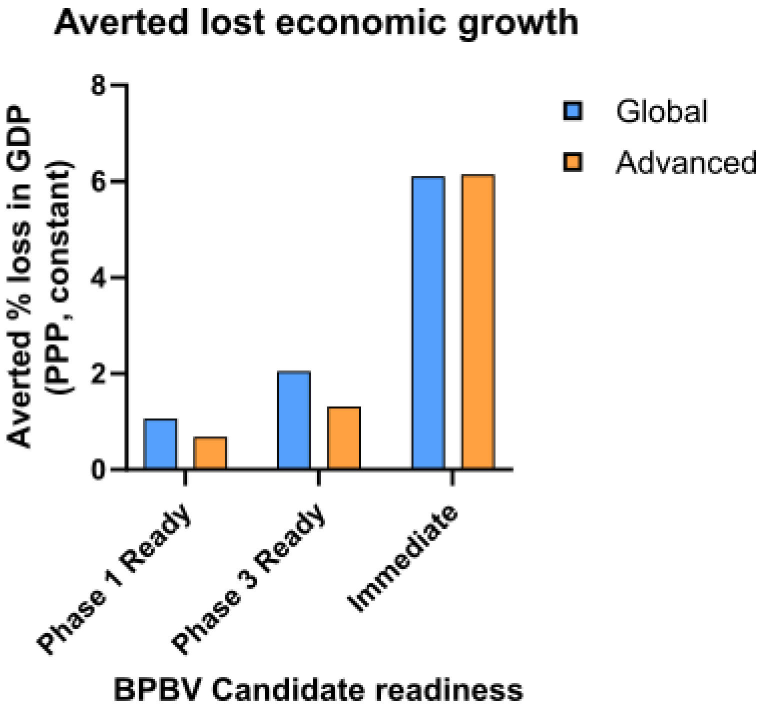


Supplementary Figure 6. Potential BPBV impact on GDP during the COVID-19 pandemic

Immediate readiness = GDP (PPP, constant prices) grows at 2015-2019 average; Phase 3 ready = 2020 GDP change + 2.5 quarters of average growth; Phase 1 ready = 2020 GDP change + 1.3 quarters of average growth. Averted loss in global GDP indicates the relative difference between the estimated GDP growth of each scenario and the observed GDP growth during 2020. Modelling for global (blue) and advanced economies only (orange) displayed.

### Statistical analysis

Two-tailed Mann–Whitney U tests were carried out for all neutralising antibody group comparisons using GraphPad Prism 10.4.

**Supplementary Table 1.** Statistical comparisons (p values) between vaccine groups displayed in Figure 2, sarbecovirus pseudotype neutralisation data. Comparisons between vaccines and the naïve (vehicle) group, as well as the DVX-BPBV against individual vaccines are displayed.

| S_T2_20 vs. Vehicle | | | S_T2_35 vs. Vehicle | | | DVX-BPBV vs. Vehicle | | |
|---|---|---|---|---|---|---|---|---|
| Antigen | | *p* | Antigen | | *p* | Antigen | | *p* |
| SARS-CoV | **** | <0.0001 | SARS-CoV | ns | >0.9999 | SARS-CoV | **** | <0.0001 |
| WIV16 | **** | <0.0001 | WIV16 | ns | 0.0932 | WIV16 | **** | <0.0001 |
| RaTG13 | **** | <0.0001 | RaTG13 | **** | <0.0001 | RaTG13 | **** | <0.0001 |
| Delta | *** | 0.0003 | Delta | **** | <0.0001 | Delta | **** | <0.0001 |
| BA.1 | * | 0.0137 | BA.1 | *** | 0.0003 | BA.1 | **** | <0.0001 |
| BA.2.75 | ** | 0.0013 | BA.2.75 | **** | <0.0001 | BA.2.75 | **** | <0.0001 |
| XBB.1.5 | ns | >0.9999 | XBB.1.5 | ns | >0.9999 | XBB.1.5 | ns | >0.9999 |
| JN.1 | ns | 0.8513 | JN.1 | ns | 0.4457 | JN.1 | ns | 0.5117 |
| BA.2.86 | ns | 0.155 | BA.2.86 | ns | 0.5901 | BA.2.86 | *** | 0.0005 |

**Supplementary Table 2.** Statistical comparisons (p values) between vaccine groups displayed in Figure 2, merbecovirus pseudotype neutralisation data. Comparisons between vaccines and the naïve (vehicle) group, as well as the DVX-BPBV against individual vaccines are displayed.

| S_T2_37 vs. Vehicle | | | DVX-BPBV vs. Vehicle | | | DVX-BPBV vs. S_T2_37 | | |
|---|---|---|---|---|---|---|---|---|
| Antigen | | *p* | Antigen | | *p* | Antigen | | *p* |
| MERS-CoV | **** | <0.0001 | MERS-CoV | **** | <0.0001 | MERS-CoV | ns | >0.9999 |
| African | **** | <0.0001 | African | **** | <0.0001 | African | ns | >0.9999 |
| Arabian | **** | <0.0001 | Arabian | **** | <0.0001 | Arabian | ns | 0.3665 |

**Supplementary Table 3.** Statistical comparisons (p values) between vaccine groups displayed in Figure 2B and 2D, sarbecovirus pseudotype neutralisation data. Comparisons between Hartley guinea pigs immunised with S_T2_20 and S_T2_35, and mice immunised with S_T2_20 or S_T2_35 alone.

| Antigen | S_T2_20+S_T2_35 vs. S_T2_20 | | S_T2_20 + S_T2_35 vs. S_T2_35 | |
|---|---|---|---|---|
| SARS-CoV | * | 0.0415 | **** | <0.0001 |
| WIV16 | *** | 0.0001 | **** | <0.0001 |
| RaTG13 | ** | 0.008 | *** | 0.0008 |
| Delta | *** | 0.0008 | *** | 0.0008 |
| BA.1 | *** | 0.0001 | ns | 0.5147 |
| BA.2.75 | *** | 0.0001 | ** | 0.008 |
| XBB.1.5 | **** | <0.0001 | **** | <0.0001 |
| JN.1 | *** | 0.0001 | *** | 0.0001 |
| BA.2.86 | *** | 0.0001 | *** | 0.0001 |

**Supplementary Table 4.** Group organisation (n=12 per group) for vaccinated (2 doses total with 3-week interval) mice challenged with SARS-CoV (A), SARS-CoV-2 Delta (B) and MERS-CoV (C). Mice receiving single antigen doses received 10µg total mRNA, while DVX-BPBV immunised mice received 20µg total mRNA (5µg per antigen).

| **Arm A (SARS-CoV) Group:** | **Vaccine** | **Dose per immunisation** | **Mice (genetic background)** |
|---|---|---|---|
| 1 | Vehicle | N/A | BALB/c |
| 2 | DVX-BPBV | 20 µg | |
| 3 | S_T2_20 | 10 µg | |
| 4 | N_T2_2 | 10 µg | |
| **Arm B (SARS-CoV-2 Delta) Group:** | **Vaccine** | **Dose** | **Mice** |
| 1 | Vehicle | N/A | B6.Cg-Tg(K18-ACE2)2Prlmn/J |
| 2 | DVX-BPBV | 20 µg | |
| 3 | S_T2_20 | 10 µg | |
| 4 | S_T2_35 | 10 µg | |
| 5 | N_T2_2 | 10 µg | |
| **Arm C (MERS-CoV) Group:** | **Vaccine** | **Dose** | **Mice** |
| 1 | Vehicle | N/A | huDPP4 |
| 2 | DVX-BPBV | 20 µg | |
| 3 | S_T2_37 | 10 µg | |

**Supplementary Table 5**. Viruses, mouse strains and suppliers used in animal infection studies.

| Virus | Virus supplier | Mouse | Mouse supplier |
|---|---|---|---|
| maSARS-CoV | NML (reverse genetics) | BALB/c | Charles River |
| SARS-CoV-2 Delta B.1.617.2 (POR2) | UKHSA, Porton Down | B6.Cg-Tg(K18-ACE2)2Prlmn/J | Jax |
| maMERS-CoV | NML (reverse genetics) | huDPP4 | University of Manitoba |

**Supplementary table 6:** Description of model parameters varied in dynamical compartmental modelling of mass-vaccination of high-risk populations with BPBV.

| | **Central Value** | **Sensitivity Analysis Range** | **Notes** |
|---|---|---|---|
| **BPBV efficacy against severe disease** | 75% | {50%, 75%, 95%} | BPBV efficacy against severe disease in breakthrough infections (i.e. where the BPBV fails to prevent the infection). |
| **BPBV efficacy against infection** | 35% | {0%, 35%} | BPBV efficacy against being infected. |
| **Duration of BPBV-induced immunity** | 365 days | | 365 days selected to reflect the assumption of minimal antibody waning over the period between BPBV vaccination and introduction of the SARS-CoV-2-specific vaccines at the end of 2020. |
| **Size of BPBV stockpile** | 50%% | {20-100%} | Size of the BPBV stockpile and associated level of BPBV coverage achieved in the 60+ year old population eligible to receive the vaccine. Three scenarios considered:<br>• High Coverage – 100% of 60+ years receive BPBV, irrespective of country.<br>• Moderate Coverage – 50% of 60+ years receive BPBV, irrespective of country.<br>• Variable Coverage – 20%/40%/60%/80% of 60+ years receive BPBV, depending on World Bank Income Strata (LIC/LMIC/UMIC/HIC). |
| **Speed of BPBV vaccination campaign** | Income-strata dependent | | Values derived in a World Bank Income Strata specific manner, based on empirical estimates from COVID-19 vaccination data from Our World In Data[23] |
| **Time to BPBV availability** | Immediately after global COVID-19 death toll reaches 100 deaths | {Immediate, 140 days, 250 days} | Sensitivity analysis selected to represent different levels of BPBV readiness ahead of the SARS-CoV-2 pandemic; specifically the following:<br>• Immediate Availability: BPBV has undergone all necessary approvals and evaluations to enable administration to individuals immediately. |

| | | | |
|---|---|---|---|
| | | | • Phase-3-Ready: BPBV is ready to be evaluated in Phase 3 trials immediately following the onset of the pandemic (moderate state of readiness).<br>• Phase-1-Ready: Phase 1 trials for BPBV are yet to be completed at the onset of the pandemic (minimal state of readiness). |

### Summary of economical modelling (Airfinity)

The 5-year pre-COVID-19 pandemic average (2015-2019) percentage growth of real (constant prices) GDP at purchasing power parity (PPP) per annum and per quarter were calculated using data obtained from the IMF. GDP change due to the COVID-19 pandemic was defined as the observed real GDP (PPP) change in 2020. For expected GDP change under the “immediate readiness” scenario, the average pre-COVID-19 annual growth was applied. For the other scenarios expected growth was calculated as GDP change during the pandemic plus normal growth applied on a pro-rata basis of how long left in a year from time of anticipated vaccine rollout - 225 days (2.5 quarters) for “Phase 3 ready” and 115 days (1.3 quarters). Averted % loss in GDP was calculated as the difference between observed 2020 growth and expected growth from each scenario.
